# Is exposure to pesticides associated with biological aging? A systematic review and meta-analysis

**DOI:** 10.1101/2024.04.04.24305319

**Authors:** Shanshan Zuo, Vidhya Sasitharan, Gian Luca Di Tanna, Judith M. Vonk, Maaike De Vries, Moustafa Sherif, Balázs Ádám, Juan Carlos Rivillas, Valentina Gallo

**Author notes:** **Corresponding author:** Shanshan Zuo Department of Sustainable Health Campus Fryslân University of Groningen Wirdumerdijk 34 8911 CE Leeuwarden The Netherlands.

## Abstract

**Objective:** Exposure to pesticides is a risk factor for various diseases, yet its association with biological aging remains unclear. We aimed to systematically investigate the relationship between pesticide exposure and biological aging.

**Methods:** PubMed, Embase and Web of Science were searched from inception to August 2023. Observational studies investigating the association between pesticide exposure and biomarkers of biological aging were included. Three-level random-effect meta-analysis was used to synthesize the data. Risk of bias was assessed by the Newcastle-Ottawa Scale.

**Results:** Twenty studies evaluating the associations between pesticide exposure and biomarkers of biological aging in 10,368 individuals were included. Sixteen reported telomere length and four reported epigenetic clocks. Meta-analysis showed no statistically significant associations between pesticide exposure and the Hannum clock (pooled β = 0.27; 95%CI: -0.25, 0.79), or telomere length (pooled Hedges’g = -0.46; 95%CI: -1.10, 0.19). However, the opposite direction of effect for the two outcomes showed an indication of possible accelerated biological aging. After removal of influential effect sizes or low-quality studies, shorter telomere length was found in higher-exposed populations.

**Conclusion:** The existing evidence for associations between pesticide exposure and biological aging is limited due to the scarcity of studies on epigenetic clocks and the substantial heterogeneity across studies examining telomere length. High-quality studies incorporating more biomarkers of biological aging, focusing more on active chemical ingredients of pesticides and accounting for potential confounders are needed to enhance our understanding of the impact of pesticides on biological aging.

## 1. Introduction

Pesticides are defined as a class of chemical compounds used to manage and control pests (Kim et al., 2017). Based on their target, pesticides are categorized mainly into herbicides, insecticides, fungicides and rodenticides, and are widely used globally to protect crops and improve productivity. Most of the pesticides applied contaminate the soil, evaporate into the atmosphere, infiltrate the water flow, and eventually enter the food chain (Sun et al., 2018). Therefore, pesticides are considered ubiquitous in the environment. People may be occupationally exposed to pesticides when farming, applying pesticides or manufacturing them (Figueiredo et al., 2022). In addition, co-habitants of farmers, residents living nearby farmland, and those who apply pesticides in the household can also be exposed (Bloem & Boonstra, 2023; Galdiano et al., 2021; Stanganelli et al., 2020).

Although acute non-intentional poisoning incidents have become infrequent, long-term and low-dose exposure to pesticides has various adverse effects to human health including neurologic diseases (Bemanalizadeh et al., 2022; Yan et al., 2016), cancers (Togawa et al., 2021), birth defects (Enyew et al., 2023) and diabetes (Jia et al., 2023). Moreover, evidence from longitudinal population-based studies indicates that exposure to pesticides is associated with a higher risk of all-cause mortality (Bao et al., 2020; Di et al., 2023).

Aging is characterized by the physiological deterioration of the human body over time (López-Otín et al., 2023). The aging process occurs alongside elevated risks of many chronic diseases and mortality, however there is a substantial between-person variation in the pace at which aging takes place (Fransquet et al., 2019). Biological age has emerged as a new measure for assessing differences in aging rates. It represents the overall aging process within bodies, which can be different from the chronological age. The difference between chronological and biological age can be quantified giving a measure of accelerated aging (Nakamura & Miyao, 2003). In recent years, various biological aging measures have been proposed and utilized in studies, including telomere length, DNA methylation estimator of telomere length (DNAmTL), epigenetic clocks including Horvath, Hannum, PhenoAge, GrimAge, DunedinPoAm and DunedinPACE (Horvath, 2013; Levine et al., 2018; Q. Wang et al., 2023), the Klemera and Doubal method (KDM), homeostatic dysregulation (HD), allostatic load (AL) and the biological health score (BHS) (Mak et al., 2023).

Numerous lifestyle factors, concurrent diseases or morbidities, and environmental exposures including exposure to pesticides were found to be associated with accelerated or decelerated aging (Oblak et al., 2021) and with mortality rates (Fransquet et al., 2019). Several epidemiological studies reported associations between pesticide exposure and biomarkers of biological aging (Cosemans et al., 2022; Lind et al., 2018). However, the evidence available is not consistent and difficult to appraise at glance, given the numerous markers available for measuring biological aging. To date, no attempts were made to capture the association between pesticide exposure and biological aging in a systematic way. Therefore, the objective of this study is to systematically review and meta-analyse the evidence on the association between exposure to pesticides and biological aging.

## 2. Methods

This systematic review and meta-analysis was reported in compliance with the Preferred Reporting Items for Systematic Reviews and Meta-Analyses (PRISMA) reporting guidelines (Page et al., 2021). The protocol was registered in PROSPERO (CRD42023392845).

### 2.1. Eligibility criteria

Inclusion criteria were applied according to PECO (Population, Exposure, Comparator, Outcome) statement (Table 1). Epidemiological studies using cohort, case-control, and cross-sectional designs and published in English were included. Related systematic reviews were not included but used to scan for references. Ecological studies, methodological papers, and conference abstracts were excluded.

**Table 1.**
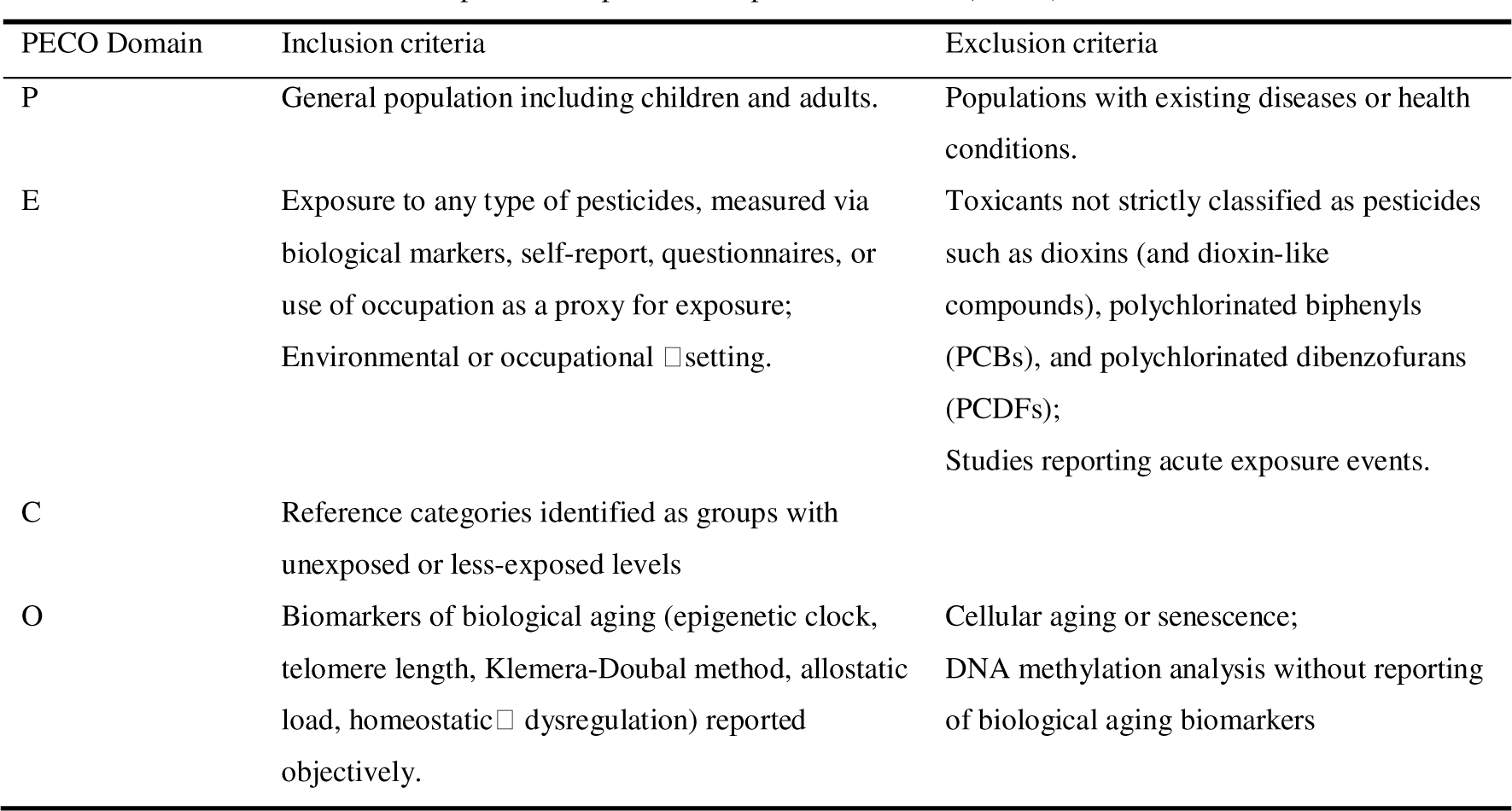
Population-Exposure-Comparator-Outcome (PECO) statement.

| PECO Domain | Inclusion criteria | Exclusion criteria |
| --- | --- | --- |
| P | General population including children and adults. | Populations with existing diseases or health conditions. |
| E | Exposure to any type of pesticides, measured via biological markers, self-report, questionnaires, or use of occupation as a proxy for exposure; Environmental or occupational □ setting. | Toxicants not strictly classified as pesticides such as dioxins (and dioxin-like compounds), polychlorinated biphenyls (PCBs), and polychlorinated dibenzofurans (PCDFs); Studies reporting acute exposure events. |
| C | Reference categories identified as groups with unexposed or less-exposed levels |  |
| O | Biomarkers of biological aging (epigenetic clock, telomere length, Klemara-Doubal method, allostatic load, homeostatic □ dysregulation) reported objectively. | Cellular aging or senescence; DNA methylation analysis without reporting of biological aging biomarkers |

### 2.2. Search strategy

Three databases (PubMed, EMBASE, Web of Science) were searched for studies published from inception to August 2023. The search strategy was developed for each database, utilizing Emtree in Embase and combining free text and MeSH terms in PubMed for retrieval. All pesticides, categorized by their targets (pesticide, insecticide, herbicide, fumigant, bactericide, and rodenticide), were searched across the databases. Outcomes comprised epigenetic clocks, telomere length, Klemera-Doubal Method, homeostatic dysregulation, allostatic load and biological health score, along with their broad categories including biological aging, accelerated aging and epigenetic age. Detailed search terms were reported in Table A. 2. Reference lists of included papers and relevant reviews were manually scanned for additional publications.

### 2.3. Study selection

References were de-duplicated in Endnote software X9. All titles and abstracts were screened for relevance then full-text examination was conducted independently by two authors (SZ and VG). Any discrepancies were resolved by discussion. The following information from the included articles was extracted: first author, year of publication, country, study design, sample size, age and gender of participants, type of pesticide(s), exposure setting and assessment, methods for biological aging estimation, statistical model, covariates adjustment, main results including maximally adjusted effect sizes along with standard error (SE) or 95% confidence intervals (CI).

For studies reporting effect sizes between exposed and non-exposed groups, sample size, mean and standard deviation (SD) of the biomarkers were retrieved from each group. When biomarker data were reported as the median and interquartile range (IQR), they were converted to mean and SD (Wan et al., 2014). If results of multiple independent subgroups were reported in one article, they were merged into a single group (Higgins et al., 2019).

If data were only presented in graphs, WebPlotDigitizer (https://automeris.io/WebPlotDigitizer/, Version 4.7) was used to extract numerical values. When studies had overlapping populations and information, the one with larger sample size or provided more information (SD, 95%CI or p-value) was selected. Corresponding authors were contacted for missing data.

### 2.4. Risk of bias assessment

Two reviewers (SZ and VS) independently assessed the risk of bias of included studies by using the Newcastle-Ottawa Scale (NOS) for cohort, case-control studies (https://www.ohri.ca/programs/clinical_epidemiology/oxford.asp), and the adapted version for cross-sectional studies (Herzog et al., 2013). A score of more than 7 was considered high quality, a score of 6–7 was considered moderate quality and a score of less than 6 was considered low quality. The higher the score, the lower the risk of bias. Any disagreements in scoring between two reviewers were resolved by the third reviewer (VG).

### 2.5. Synthesis of results

From the initial search, telomere length and epigenetic clock estimates were the only two biomarkers of biological aging identified. Synthesis of results encompassed meta-analysis, sensitivity analysis and moderator analysis. For studies that could not be quantitatively analysed in the meta-analysis, the results were summarized in a narrative form.

#### 2.5.1. Meta-analysis

Meta-analysis was performed where comparable data were available in at least two studies. To account for the multiple dependent effect sizes due to more than one pesticide reported within one study, a three-level random-effects model was employed (Harrer et al., 2021). Contrary to the traditional univariate meta-analytic approach, which only considers sampling variance of the extracted effect sizes and variance between studies, the three-level meta-analysis additionally models the variation of effect sizes within studies (Assink & Wibbelink, 2016; Cheung, 2014).

All studies using epigenetic clock as an outcome reported the linear relationship between pesticide exposure as ln-transformed continuous variable and epigenetic clock. In studies which used telomere length as an outcome, various exposure assessment methods led to reporting of different types of effect sizes (i.e., β coefficient, mean difference). The corrected standardized mean difference (Hedges’ g) on the raw scale was chosen as the effect size for the meta-analysis, representing the mean difference in telomere length between individuals with higher and lower exposure to pesticides. Positive values indicated longer telomere length in individuals with higher exposure compared to those with lower exposure, while negative values suggested shorter telomere length. The application of Hedges’ g allowed the maximisation of inclusion of the results in the meta-analysis; the majority of studies reported telomere length in exposed and unexposed groups. Moreover, effect size transformation methods were available to convert β coefficients from linear regression models with binary and continuous exposure variables into Hedges’ g. Formulas for calculation and transformation were presented in Appendix B.

Heterogeneity was estimated by the *I^2^* statistic using the var.comp function in *demeta* (Cheung, 2014). The total amount of heterogeneity was decomposed into: a) sampling variance at level 1, b) within-study heterogeneity at level 2 (*I^2^_within_*), and c) between-study heterogeneity at level 3 (*I^2^_between_*). In addition, statistical significance of heterogeneity was tested using Cochran’s Q statistic.

#### 2.5.2. Sensitivity analysis

Outlier detection and influence diagnostics were performed to elucidate the source of heterogeneity and test the robustness of the pooled effect estimate. Effect sizes falling below the first quartile or above the third quartile of the interquartile range (IQR) were considered outliers (Gómez Penedo & Flückiger, 2023). Effect sizes were considered influential if their Cook’s distance exceeded three times the mean Cook’s distance across included studies (Ellis et al., 2023). Sensitivity analysis was carried out by removing outlier effect sizes, influential effect sizes, low-quality studies, and effect sizes with transformations.

#### 2.5.3. Moderator analysis

Additional moderator analysis was performed using omnibus tests to explore potential sources of heterogeneity across the following moderators: chemical class of pesticide (organophosphate or organochlorine), gender (male or female), exposure setting (occupational or environmental) and expression of telomere length (T/S ratio, bp or kb/genome) (Assink & Wibbelink, 2016).

#### 2.5.4. Publication bias

Publication bias were assessed through visual inspection of funnel plots in combination with the Egger’s test. All statistical analyses were conducted using the *metafor* and *dmeta* packages of R statistical software (version 4.3.0).

## 3. Results

### 3.1. Study selection

The initial search retrieved 2079 records; after 540 duplicates were removed, 1539 records were screened by title and abstract, and 44 articles were selected for full-text eligibility assessment. Among these, 24 articles were excluded (reasons for exclusion were in Fig. 1), yielding a total of 20 eligible articles for the systematic review (Ali et al., 2023; Andreotti et al., 2015; Cosemans et al., 2022; de Oliveira et al., 2019; de Prado-Bert et al., 2021; dos Santos et al., 2022; Duan et al., 2017; Guzzardi et al., 2016; Hoang et al., 2021; Hou et al., 2013; Kahl, da Silva, et al., 2018; Kahl, Dhillon, Fenech, et al., 2018; Kahl, Dhillon, Simon, et al., 2018; Kahl et al., 2016; Karimi et al., 2020; Lind et al., 2018; Lucia et al., 2022; Ock et al., 2020; Saad-Hussein et al., 2019; Shin et al., 2010). Two additional articles were identified through reference lists of selected systematic reviews (Passos et al., 2022) (Fig. 1). Overall, 17 studies were included in meta-analyses while 3 studies were included in the narrative synthesis only.

**Fig. 1.**
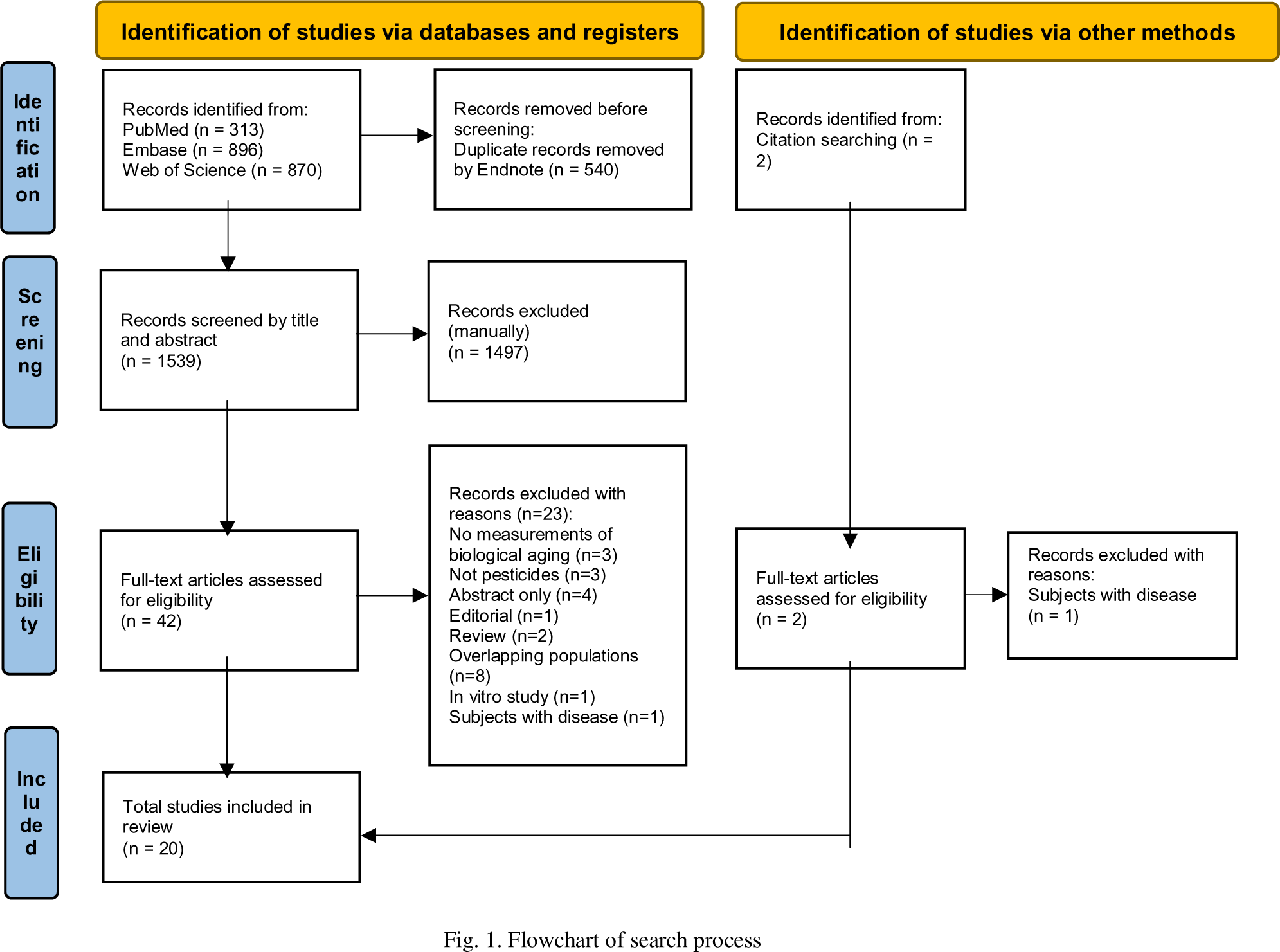
Flowchart of search process

Despite the overlapping population between (Hou et al., 2013) and (Andreotti et al., 2015), they were treated as two separate studies and included in the systematic review since the size of overlap was small (40/1234 and 40/568).

### 3.2. Characteristics of the studies

Summary characteristics of the studies were described in Table 2 and Table 3. Twenty studies evaluating the associations between pesticide exposure and two biomarkers of biological aging in 10368 individuals aged between 7 to 92 years (6647 females and 3721 males) were included. Sixteen studies reported telomere length and four studies reported epigenetic clocks. The vast majority were cross-sectional studies, with only 3 studies using a cohort design. The studies were carried out in South America (n = 6), North America (n = 5), Europe (n = 4), Asia (n = 4) and Africa (n = 1). More studies assessed occupational exposures (55%) rather than environmental exposures (40%), while the sample in one study was composed of occupational exposed and non-occupational exposed (Ali et al., 2023). The included studies primarily assessed exposure through the concentration of pesticide metabolites in biological samples (blood/urine). Other approaches included using occupational exposure as a proxy for exposure and self-reported exposure by questionnaires.

**Table 2.** Characteristics of included studies on pesticide exposure and epigenetic clocks.

| Author, year | Country | Study population | N (female %) | Study design | Pesticides | Exposure setting | Exposure assessment | Clocks | Sample, platform | Adjustments |
| --- | --- | --- | --- | --- | --- | --- | --- | --- | --- | --- |
| De Prado-Bert, et al., 2021 | Multi-countries across Europe: UK, France, Spain, Lithuania, Norway and Greece | Children from HELIX project, six longitudinal population-based birth cohorts across Europe | 1173 (45%) | Prenatal exposome: cohort, childhood exposome: cross-sectional | DDE, DDT, HCB, DEP, DETP, DMP, DMTP, DMDTP (only in childhood) | Environmental | Blood | Horvath's Skin and Blood | Blood, 450K | Child's sex, cohort, self-reported maternal education, self-reported ancestry, maternal age at conception |
| Hoang, et al., 2021 | USA | Licensed pesticide applicators from Agricultural Health Study | 1170 (0%) | Cross-Sectional | Dicamba, Picloram, Mesotrione, Acetochlor, Metolachlor, Glyphosate, 2,4-D, Atrazine, Malathion, Aldrin, Chlordane, DDT, Dieldrin, Heptachlor, Lindane, Toxaphene | Occupational | Questionnaire | Horvath, Hannum, Levine, Horvath skin and blood, GrimAge | Blood, 450K | Asthma status, age at blood collection, pack-years, smoking, state of residence, cell types |
| Lind, et al., 2018 | Sweden | Community-dwelling elderly from Prospective Study of the Vasculature in Uppsala Seniors (PIVUS) | 967 (50%) | Cross-Sectional | p,p'-DDE, HCB, TNC | Environmental | Blood | Hannum | Blood, 450K | Sex, education level, exercise habits, smoking, energy and alcohol consumption and BMI |
| Lucia, et al., 2022 | USA | Postmenopausal women residing in southern California | 392 (100%) | Cross-Sectional | Glyphosate, AMPA | Environmental | Urine | Horvath, Hannum, Levine, DunedinPoAm | Blood, 450K | Urinary creatinine concentration, age, race/ethnicity, BMI, smoking status, alcohol consumption, organic eating, diet quality, batch, position on chip and WBC type proportions |

HCB: hexachlorobenzene; TNC: transnonachlor; DMP: dimethylphosphate; DEP: diethylphosphate; DMTP: dimethylthiophosphate; DETP: diethylthiophosphate; DMDTP: dimethyldithiophosphate; DEDTP: diethyldithiophosphate; TCPY: 3,5,6-trichloro-2-pyridinol

**Table 3.**
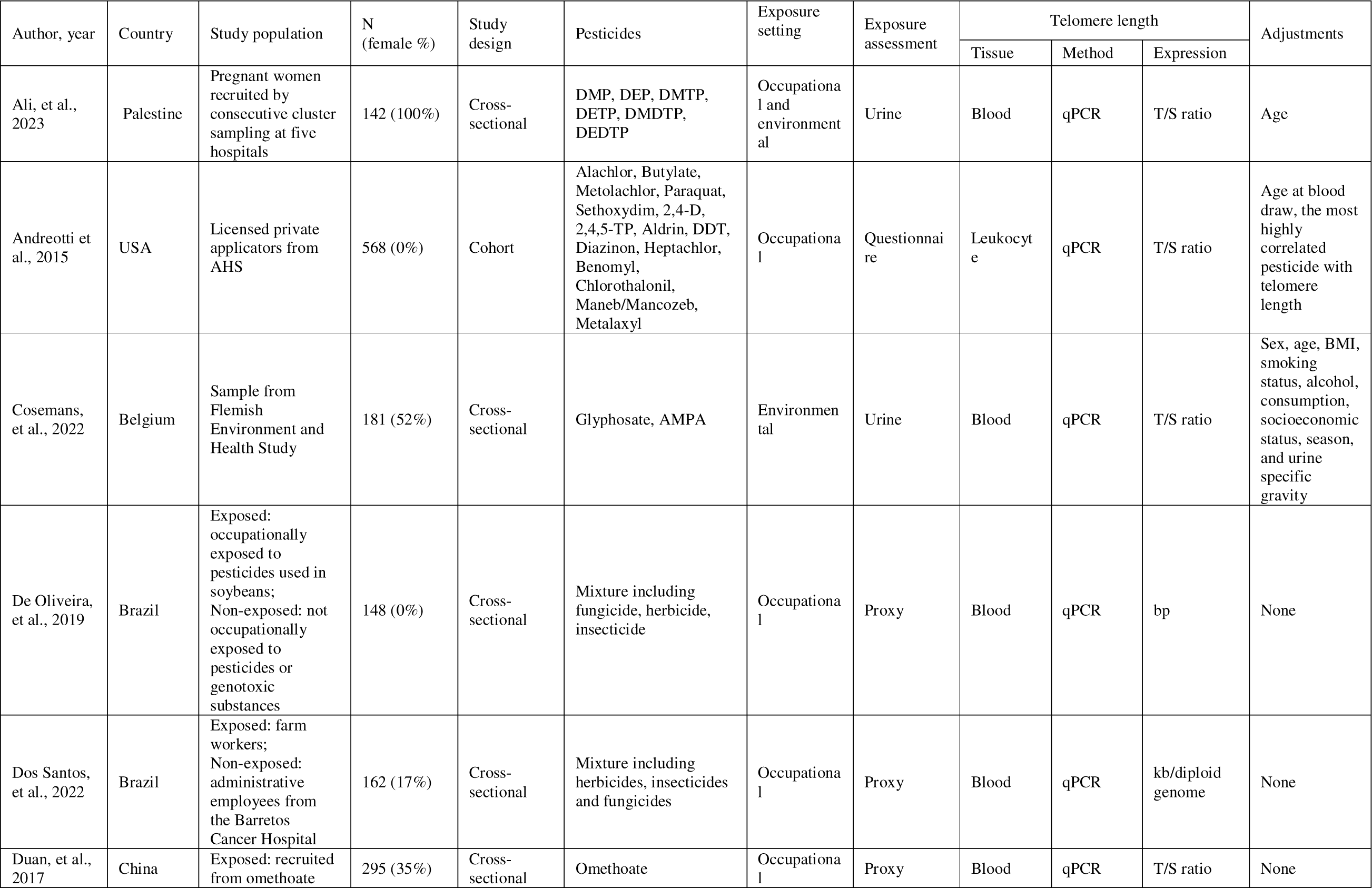
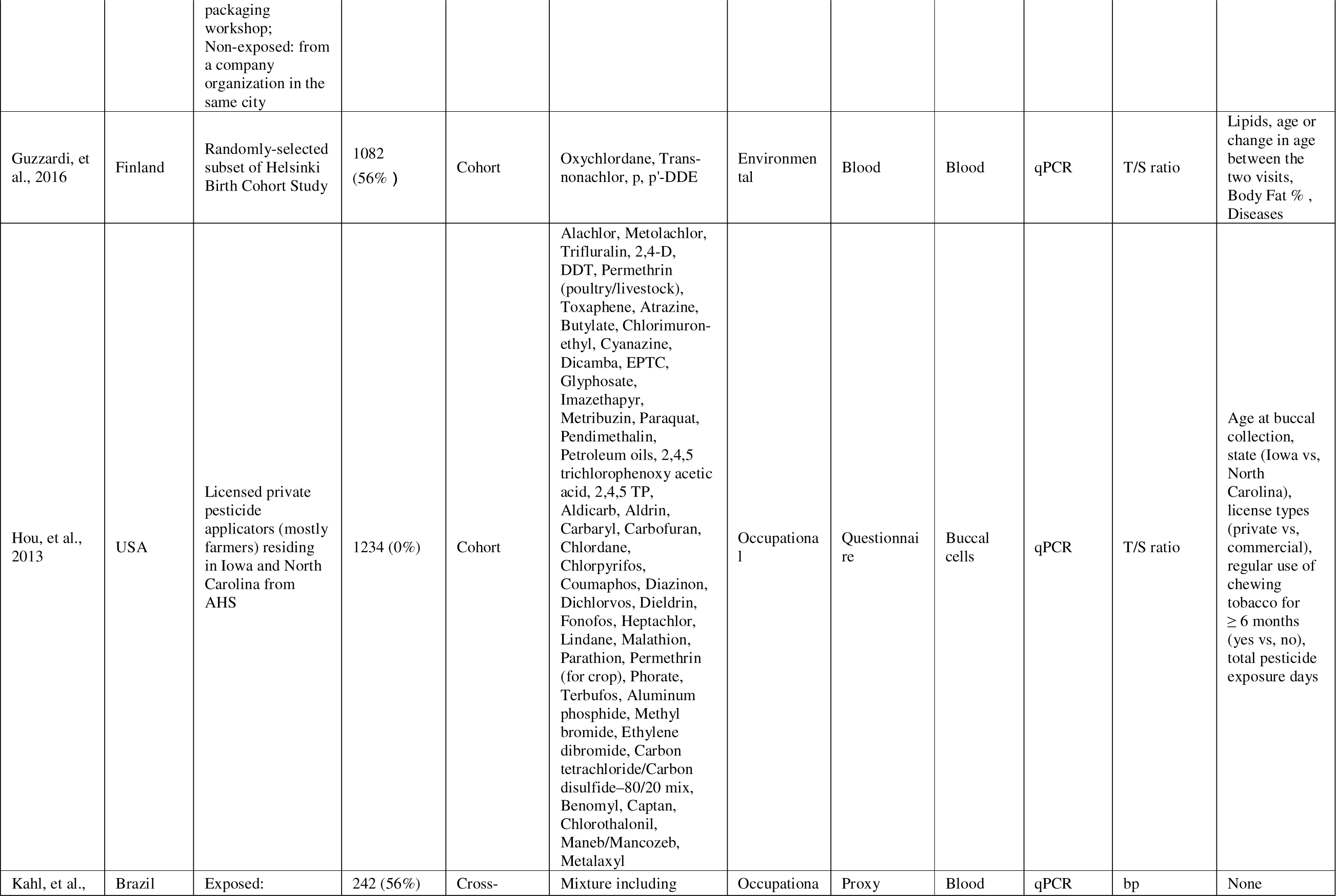
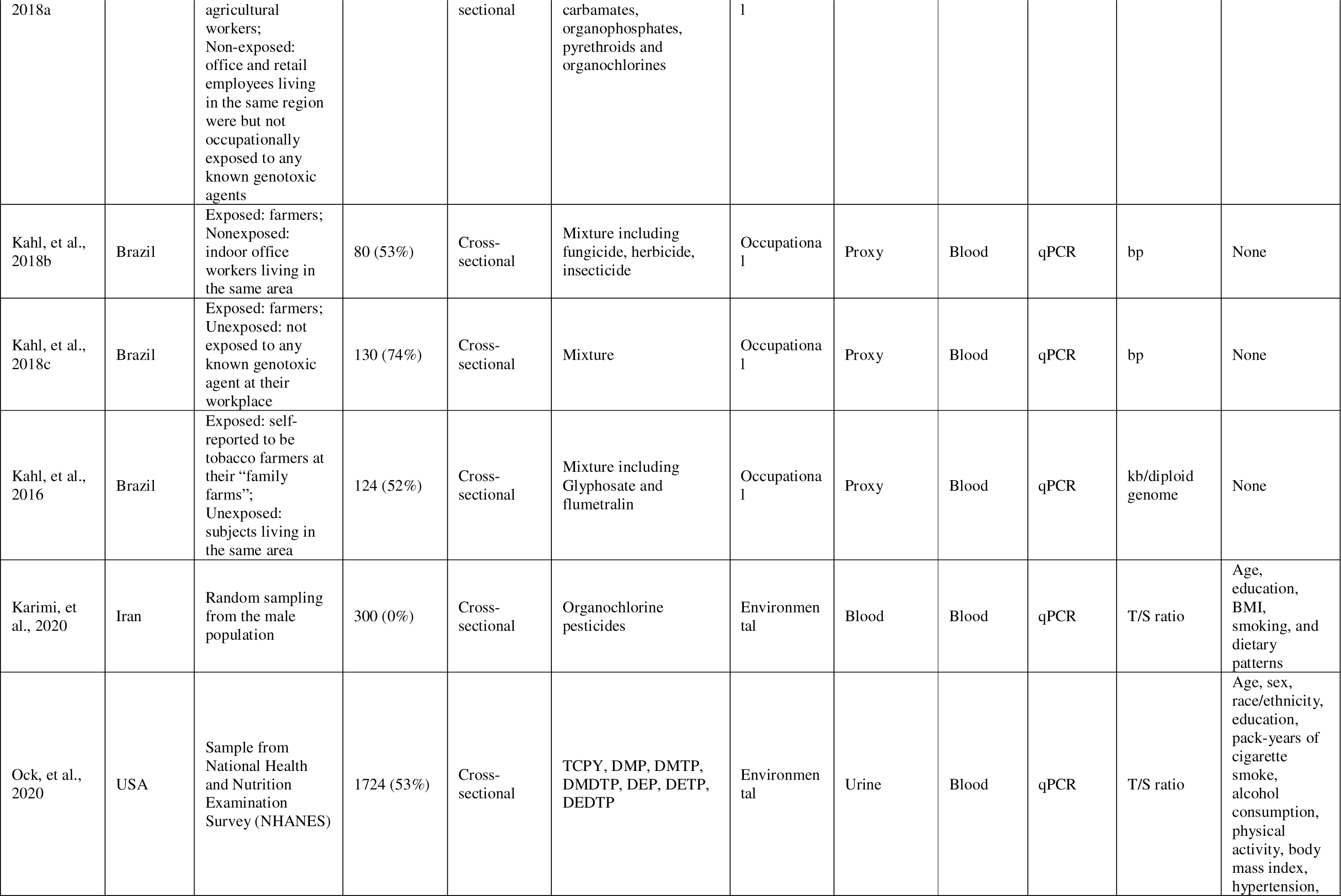
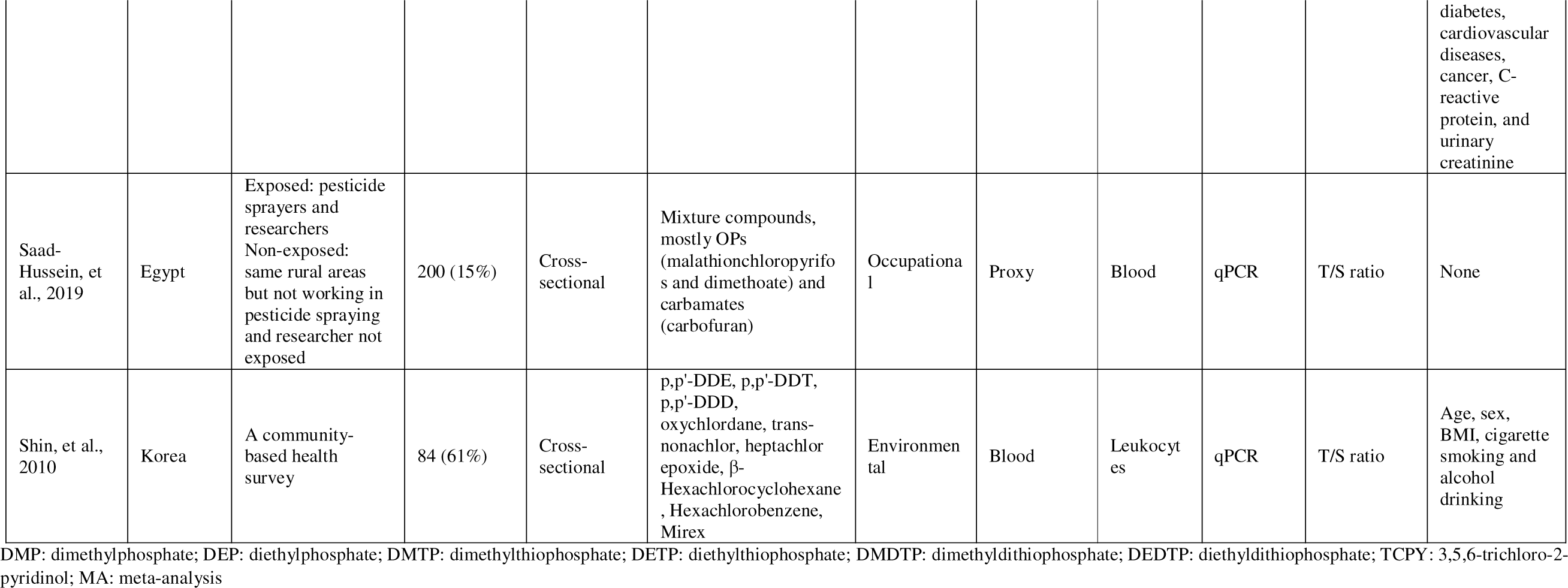
Characteristics of included studies pesticide exposure and telomere length.

### 3.3. Risk of bias

The quality scores of the selected studies were in the range of 5-9 points (Table A. 5 and Table A. 6). A total of 9 studies were rated high quality, considering adjustments for important confounding variables, and assessing exposure in biological samples or through validated tools. Eight studies were considered as moderate quality, and three were categorized as low quality due to insufficient control of confounding factors or because they relied on occupation as a proxy for exposure, lacking precise or validated tools for exposure assessment.

### 3.4. Associations between pesticide exposure and epigenetic age

A total of four studies assessed the association between pesticide exposure and epigenetic clocks (Table 2 and Table A. 3). The majority used a cross-sectional design, with only one study having both a longitudinal and a cross-sectional component, investigating prenatal (cohort) and childhood (cross-sectional) pesticide exposure in relation to children’s epigenetic age (de Prado-Bert et al., 2021). The total sample size of the included studies was 3672, comprising 2267 males and 1405 females. Three studies focused on middle-aged and elderly individuals, with ages ranging from 45 to 70 years. One study was conducted among children, with an average age of 7 years.

A total of 25 metabolites from 6 chemical categories were reported, with organochlorine and organophosphate pesticides being the most analysed. One study included a wide range of pesticides including chlorophenoxy (Dicamba, 2,4-D), chloroacetanilides (Acetochlor, Metolachlor), Pyridine (Picloram) and Trazines (Atrazine) (Hoang et al., 2021). Most studies focused on more than one epigenetic clock, including Horvath (Hoang et al., 2021; Lucia et al., 2022), Hannum (Hoang et al., 2021; Lind et al., 2018; Lucia et al., 2022), PhenoAge (Hoang et al., 2021; Lucia et al., 2022), DunedinPoAm (Lucia et al., 2022), GrimAge (Hoang et al., 2021), and Horvath Skin and Blood (de Prado-Bert et al., 2021; Hoang et al., 2021). Among these, the Hannum clock was the most frequently utilized, being employed in three studies.

Three studies reported positive significant associations between metabolites of four pesticides (p,p’-DDE, DDT, TNC, and AMPA) and accelerated epigenetic aging (Hoang et al., 2021; Lind et al., 2018; Lucia et al., 2022). On the contrary, one study reported a negative association between dimethyldithiophosphate (DMDTP) and epigenetic age; they interpreted this as the result of higher intake of fruits and vegetables; however the results did not change substantially once additionally adjusted for fruit intake, vegetable intake or urinary hippurate, a metabolite marker of fruits and vegetables (de Prado-Bert et al., 2021).

Two studies reporting five effect sizes of the association between levels of pesticide metabolites and the Hannum clock were included in the meta-analysis. Results were presented in the forest plot (Fig. 2). Overall, a positive but not significant association was found between pesticide exposure and epigenetic age (pooled β= 0.27; 95%CI: -0.25, 0.79). 23.83% of overall heterogeneity was attributable to between-study differences (*I^2^_between_*), while 42.57% was attributable to within-study differences (*I_2within_*).

**Fig. 2.**
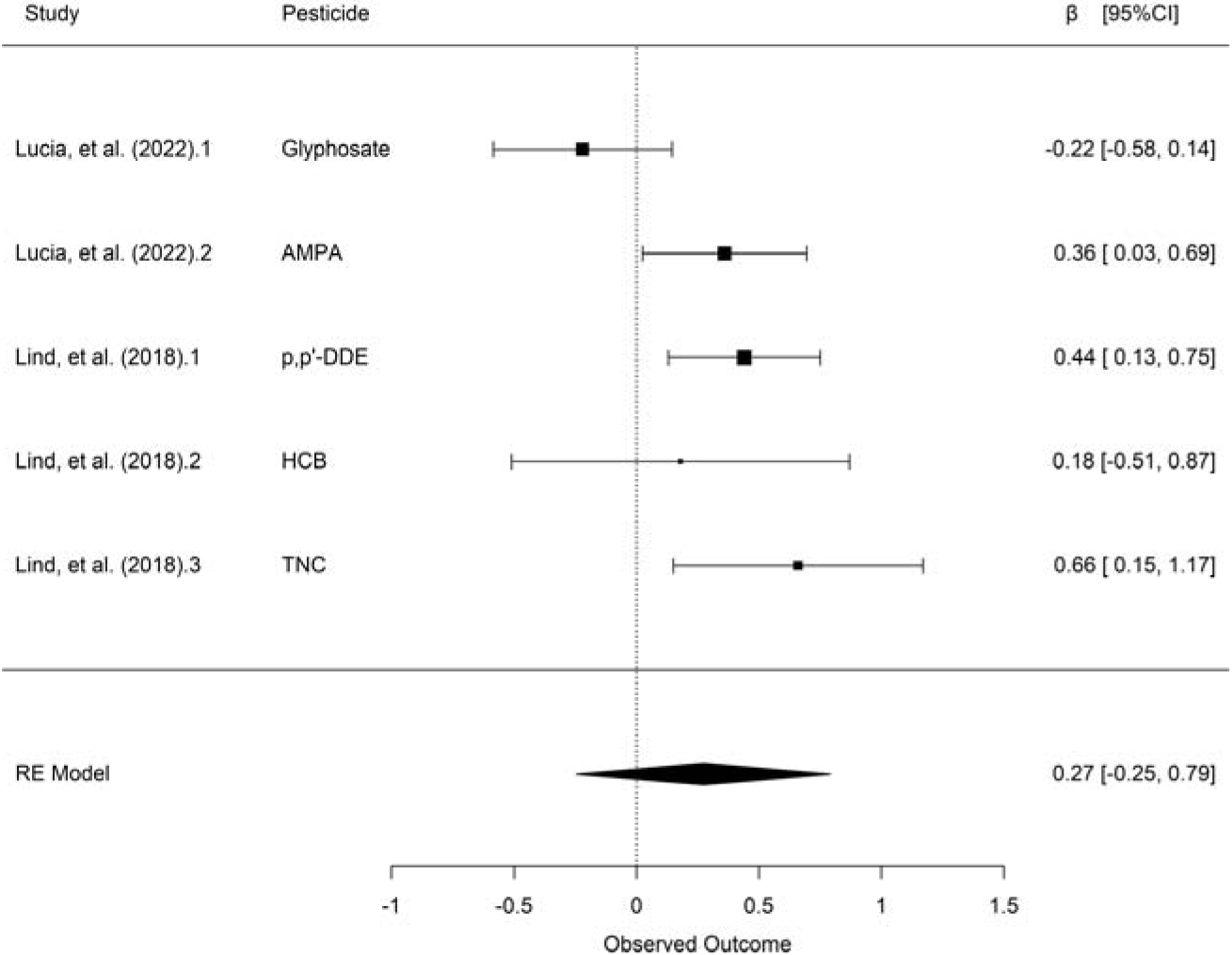
Forest plot for meta-analysis of associations between pesticide exposure (ln-transformed) and Hannum epigenetic age

### 3.5. Associations between pesticide exposure and telomere length

Sixteen studies reported the association between pesticide exposure and telomere length (Table 3). The sample sizes ranged from 80 to 1724, resulting in a total of 6696 participants, including 4380 males and 2316 females. Three studies measured exposure from blood samples (Guzzardi et al., 2016; Karimi et al., 2020; Shin et al., 2010), three from urine samples (Ali et al., 2023; Cosemans et al., 2022; Ock et al., 2020); two studies used questionnaires (Andreotti et al., 2015; Hou et al., 2013), and the remaining eight categorized participants into exposed and non-exposed groups based on their occupation and exposure history (de Oliveira et al., 2019; dos Santos et al., 2022; Duan et al., 2017; Kahl, da Silva, et al., 2018; Kahl, Dhillon, Fenech, et al., 2018; Kahl, Dhillon, Simon, et al., 2018; Kahl et al., 2016; Saad-Hussein et al., 2019).

Nine studies reported results of in total 72 specific pesticides, mainly belonging to the organochlorine and organophosphate classes, followed by carbamate. The remaining seven studies reported on pesticide mixtures. In all studies, telomere length was assessed using the quantitative polymerase chain reaction (qPCR) method. Specifically, telomere length was expressed as the ratio of telomere repeat copy to the relative number of a single copy gene (T/S ratio) in ten studies, as base pairs (bp) in four studies, and as kb/diploid genome in two studies. Overall, the association between pesticide exposure and telomere length was not consistent (Table A. 4): ten studies reported shorter telomeres and two studies reported longer telomeres (Cosemans et al., 2022; Duan et al., 2017), one study found shorter telomeres with diethylphosphate (DE) derived metabolites but not with dimethylphosphate(DM) derived metabolites (Ali et al., 2023). In one study, a negative association was found between 3,5,6-trichloro-2-pyridinol (TCPY) and telomere length, while a positive association was found for diethyl thiophosphate (DETP) (Ock et al., 2020). Furthermore, one study indicated an inverted U-shaped change in telomere length with increasing pesticide concentration (Shin et al., 2010), and two studies found no overall association between pesticide exposure and telomere length (de Oliveira et al., 2019; dos Santos et al., 2022).

One study was excluded from the meta-analysis because it reported neither standard error, confidence intervals, or p-values. Of the 15 studies included in the primary meta-analysis, 11 studies presented means of telomere length in exposed and unexposed groups. Hedges’ g was directly calculated from mean difference data. The remaining study results were converted from β coefficients from linear regression models with binary and continuous exposure variables into Hedges’ g.

Results of the meta-analysis of the association between pesticide exposure and telomere length were shown in the forest plot (Fig. 3). Overall, telomere length in higher-exposed populations was shorter compared to the lower exposed population, but the association fell short of statistical significance (pooled g= -0.46; 95%CI: -1.10, 0.19). 99.15 % of the heterogeneity is explained by between-study, 0.39 % and by within-study.

**Fig. 3.**
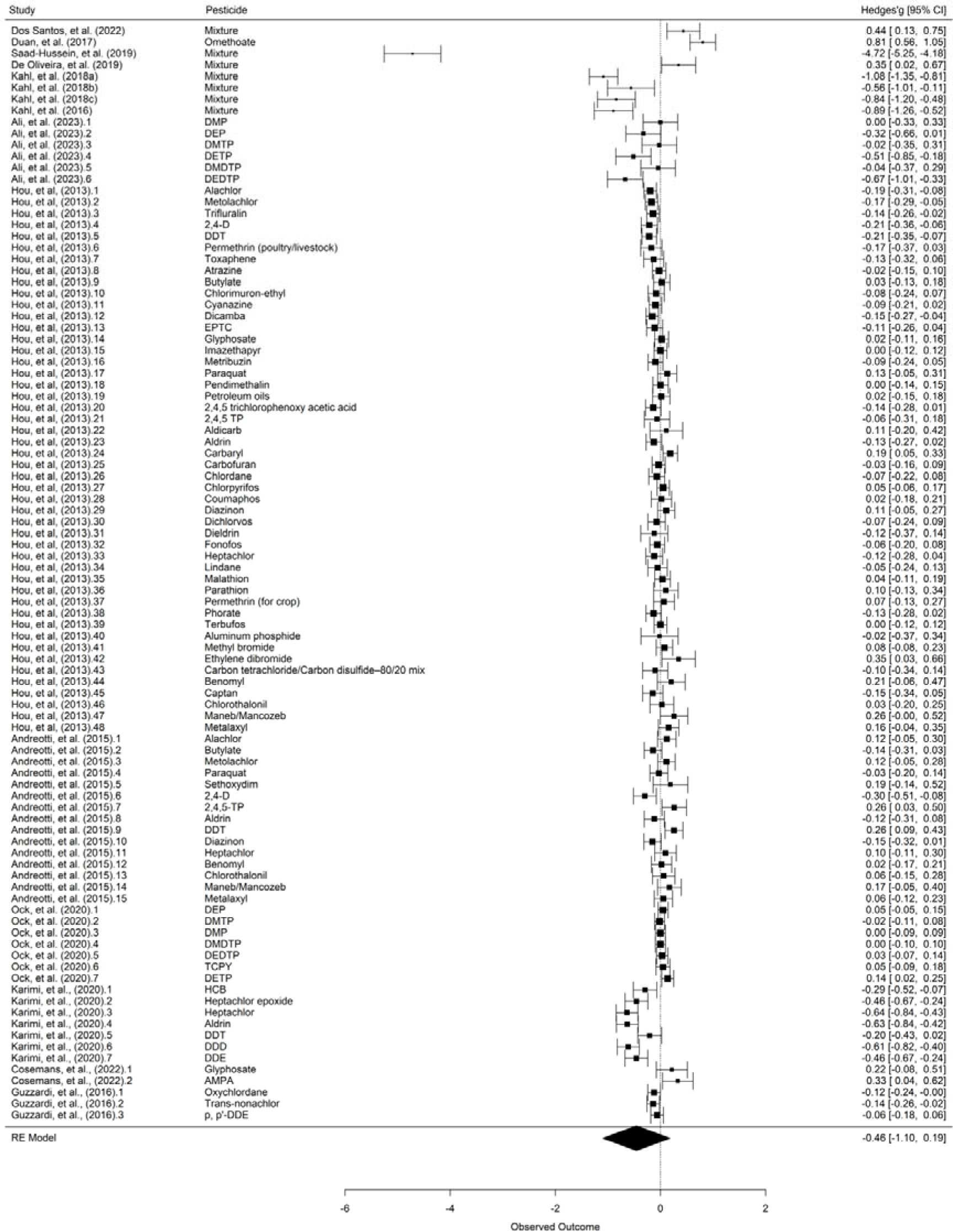
Forest plot for meta-analysis of associations between pesticide exposure and telomere lengt

#### 3.5.1. Sensitivity analysis

The sensitivity analysis showed that excluding effect sizes identified as outliers, or through transformation, did not have a strong influence on the results and the pooled effects remained negative in direction.

Notably, significant shorter telomere length in higher exposed population was found after removing the influential effect sizes (pooled g= -0.28; 95%CI: -0.54, -0.02) or low-quality studies (pooled g= - 0.29; 95%CI: -0.54, -0.01) (Table 4).

**Table 4.** Sensitivity analysis on the relationship between pesticide exposure and telomere length.

| Category | Studies (k) | Effects sizes (n) | Hedges' g | 95% CI | % Var (level 2) | % Var (level 3) |
| --- | --- | --- | --- | --- | --- | --- |
| Main analysis | 15 | 96 | -0.46 | [-1.10, 0.19] | 0.39 | 99.15 |
| Outliers removed | 8 | 82 | -0.01 | [-0.10, 0.09] | 21.46 | 52.17 |
| Influential studies removed | 12 | 93 | -0.28 | [-0.54, -0.02] | 3.10 | 93.42 |
| Low-quality studies removed | 12 | 93 | -0.29 | [-0.54, -0.01] | 2.94 | 93.76 |
| Effect sizes with transformation removed | 11 | 77 | -0.60 | [-1.48, 0.27] | 0.37 | 99.25 |
| Effect sizes with no adjustment of confounders removed | 5 | 33 | -0.06 | [-0.31, 0.19] | 8.25 | 84.05 |

#### 3.5.2. Moderator analysis

A summary of the moderator results was presented in Table 5. Chemical class of pesticide, gender, exposure setting, or expression of telomere length did not moderate the effect between pesticide exposure and telomere length.

**Table 5.** Moderator analysis on the relationship between pesticide exposure and telomere length.

| Category | Studies (k) | Effects sizes (n) | Hedges' g | 95% CI | F (df1, df2) | P | % Var (level 2) | % Var (level 3) |
| --- | --- | --- | --- | --- | --- | --- | --- | --- |
| **Pesticide class** |  |  |  |  | 0.10 (1, 47) | 0.10 | 2.49 | 92.3 |
| Organophosphate | 6 | 27 | 0.05 | [-0.20, 0.3] |  |  |  |  |
| Organochlorine | 4 | 22 | -0.02 | [-0.28, 0.23] |  |  |  |  |
| **Gender** |  |  |  |  | 0.75 (1, 82) | 0.39 | 1.97 | 96.03 |
| Female | 5 | 10 | -0.25 | [-0.75, 0.25] |  |  |  |  |
| Male | 7 | 74 | -0.39 | [-0.87, 0.08] |  |  |  |  |
| **Exposure setting** |  |  |  |  | 0.52 (1, 88) | 0.47 | 0.32 | 99.30 |
| Occupational | 10 | 71 | -0.64 | [-1.47, 0.20] |  |  |  |  |
| Environmental | 4 | 19 | -0.07 | [-1.38, 1.25] |  |  |  |  |
| **TL expression** |  |  |  |  | 0.04 (2, 93) | 0.96 | 0.34 | 99.28 |
| T/S ration | 9 | 90 | -0.48 | [-1.37, 0.42] |  |  |  |  |
| bp | 4 | 4 | -0.53 | [-1.88, 0.82] |  |  |  |  |
| kb/genome | 2 | 2 | -0.23 | [-2.14, 1.68] |  |  |  |  |

#### 3.5.3. Publication bias

The funnel plot analysis (Fig. A.1) showed the asymmetrical distribution and Egger’s test was statistically significant (P=0.0051), suggesting publication bias favoring studies with null-effect sizes.

## 4. Discussion

This is the first systematic review summarizing the current evidence on the association between pesticide exposure and biological ageing. Using a three-level meta-analysis to quantitatively synthesize data. In our review, pesticide exposure was not statistically associated with either the epigenetic clock or with telomere length. However, the opposite direction of the pooled effects for both outcomes – the positive on epigenetic age and the negative on telomere length, showed a possible indication of biological aging acceleration in relation to pesticide exposure. Importantly, after removal of influential effect sizes or low-quality studies pesticide exposure was significantly associated with shorter telomere length.

The aging process is a gradual and multifaceted phenomenon characterized by physiological deterioration at the cellular, tissue, and organ levels, resulting in an increased susceptibility to various diseases. Twelve hallmarks of aging are considered to contribute to the aging process: genomic instability, telomere attrition, epigenetic alterations, loss of proteostasis, disabled macroautophagy, deregulated nutrient sensing, mitochondrial dysfunction, cellular senescence, stem cell exhaustion, altered intercellular communication, chronic inflammation and dysbiosis (López-Otín et al., 2013).

In this systematic review, telomere length and epigenetic clocks were the only biomarkers measuring biological aging captured from existing research in relation to exposure to pesticides. Located at the termini of eukaryotic chromosomes, telomeres play a crucial role in preventing degradation during mitosis (Coltell et al., 2023). This protective function is accomplished through the addition of 5′-TTAGGG-3′ tandem repeats to the chromosome ends by telomerase, an enzyme predominantly active in germ cells, stem cells, and immortalized cells (Martens & Nawrot, 2016). Previous studies have found that shorter telomere length was associated with higher mortality (Xiong et al., 2023), development of degenerative diseases, and cardiovascular disease (X. Wang et al., 2023). In numerous epidemiological studies, it was employed as a measurement for biological aging and to predict an individual’s health condition. However, the variety of approaches used to quantify telomere length sometimes led to varying outcomes, raising concerns about its accuracy (Lin & Yan, 2005). Based on 140 CpGs and validated in leukocytes from several cohorts, Lu et al. developed a DNA methylation estimator of TL (DNAmTL), which outperformed several limitations of the classical techniques in epidemiological studies (Lu, Seeboth, et al., 2019).

The epigenetic clock was developed on large-scale genomewide DNA methylation-based epigenetic analyses using a variety of cell types and tissues and serves as another molecular marker for biological aging (Horvath, 2013). Composed of a subset of cytosinephosphate-guanine sites (CpGs), epigenetic clock-based DNA methylation age can predict chronological age and was associated with mortality and age-related morbidities (Williams et al., 2023). The first generation of epigenetic clocks, represented by Horvath’s and Hannum’s, exhibited remarkable accuracy in predicting chronological age (Zhou et al., 2022). The second generation comprises PhenoAge and GrimAge. Instead of solely focusing on the correlation with chronological age, the two clocks also incorporated phenotypic or behavioral surrogate markers that reflect age-related health, disease phenotypes, and all-cause mortality (Levine et al., 2018; Lu, Quach, et al., 2019). However, the diverse epigenetic clocks showed varying degrees of associations with risk factors and outcomes, indicating the potential to capture distinct aspects of the aging process (Oblak et al., 2021). The third generation of clocks, represented by the DunedinPACE, have been developed recently. Through using birth cohort data within-individual decline in 19 indicators of organ-system integrity across two decades to model Pace of Aging, these clocks have showed high test reliability and are considered as a complement to the second generation epigenetic clocks (Belsky et al., n.d.). Moreover, some other markers for measuring biological aging have emerged, such as the Klemera and Doubal method and Biological Health Score (BHS). These approaches measure the aging by integrating biomarkers from various systems of the body and have demonstrated high reliability in predicting biological age and mortality (K. Li et al., 2024). Current recommendations for estimating biological aging propose the necessity of a comprehensive and integrative score that incorporates both molecular biomarkers and physiological functional parameters (Khan et al., 2017).

### 4.1. Pesticides and epigenetic age

Generally, the meta-analysis showed a positive but nonsignificant association between pesticide exposure and the Hannum clock. However, the results should be interpreted with caution due to the low number of studies included in the meta-analysis, and the inability to incorporate other epigenetic clocks. On one hand, in contrast to the Hannum clock, which was developed to estimate chronological age, the second-generation clocks demonstrated superior performance in predicting age-related clinical phenotypes and all-cause mortality (McCrory et al., 2020). On the other hand, in comparison to the Horvath clock algorithm, designed as a robust multi-tissue age predictor based on DNA methylation at 353 CpG sites, the Hannum clock serves as a blood-based estimator, characterized by DNA methylation at 71 CpG sites (Fransquet et al., 2019). Across all included studies, positive statistical associations with epigenetic clocks were reported for p,p’-DDE, DDT, TNC, and AMPA. The exact biochemical mechanisms linking pesticides to accelerated aging remain uncertain. However, the connection is supported by some epigenome-wide association studies (EWAS) where differential methylation was identified for pesticides. A case-control study found correlations between organophosphate exposure and methylation levels at 70 significant CpGs, with overrepresentation of genes related to GABA-B receptor II and endothelin signaling pathways, which had impact on neurotransmitter release and inflammation, respectively (Paul et al., 2018). Another study in the Netherlands also reported differential methylation related to occupational pesticide exposure, and further found that differential methylation at specific genomic locations induced by pesticides may play a role in airway disease pathogenesis (Plaat et al., 2018). One recent study by (Hoang et al., 2021) found that glyphosate was linked to 24 differentially methylated CpG sites, while 162 CpG sites were discovered in association with eight pesticides (acetochlor, atrazine, dicamba, malathion, metolachlor, mesotrione, picloram, and heptachlor).

### 4.2. Pesticides and telomere length

A previous systematic review on occupational pesticide exposure and telomere length included a total of six studies which only reported data on exposed and non-exposed groups (Passos et al., 2022). We have now extended this including sixteen papers, including any exposure route, and including more data in the meta-analysis by using Hedges’g. Our results indicated a negative association between pesticide exposure and telomere length, which however did not reach statistical significance. Moderator analysis make no significant change but showed a concordant direction in the pooled results. The potential mechanisms through which pesticides affect telomere shortening involve mainly oxidative stress. Telomeres are extremely sensitive to damage by oxidative stress because of the high guanine content in certain telomere sequences and the deficiency in the repair of single strand breaks (Honda et al., 2001; Hou et al., 2013). In contrast, some epidemiological studies reported telomere lengthening with adverse health outcomes such as breast cancer (Y. Li & Ma, 2022), gastric cancer (Z. Wang et al., 2018), and lung cancer (Doherty et al., 2018; Tsatsakis et al., 2023). A longer telomere could result in prolonged cell survival, subsequently elevating the probability of accumulating mutations associated with cancer (Aviv et al., 2017).

Substantial heterogeneity between studies was found and this heterogeneity could not be explained after moderator analysis. It may be attributed to systematic differences in characteristics of the study populations as well as to the assessment of exposure. Among the included studies, different pesticide exposure methods were used by the individual studies, such as biological samples, questionnaires and use of occupation as a proxy for exposure. Moreover, despite the wide range of categories of pesticides studied, seven studies concentrated on a broadly defined pesticide category. Pesticides is a heterogeneous group of very different active chemical ingredients, even within their broad application group of insecticides, herbicides and fungicides, which might mix chemicals with no health effects and chemical with a rather significant health effect (Karalexi et al., 2021). In sensitivity analysis, after removing the influential studies or the low-quality studies, statistically significant associations of pesticide exposure with shorter telomere length were found, which highlighted the importance of longitudinal studies with larger sample sizes and proper adjustment for confounders in producing meaningful results. For publication bias, the funnel plot suggested that studies showing a significant negative association between pesticide exposure and telomere length could be missing, and, consequently, our effect size could be underestimated.

### 4.3. Strengths and limitations

This is the first comprehensive systematic review examining the association between pesticide exposure and biological aging. Considering that the majority of studies reported effects of more than one pesticide, a three-level meta-analytic model was employed to include all effect sizes in our analysis. This approach not only addresses the dependency of effect sizes but also ensures that all available information is preserved, allowing for the attainment of maximum statistical power (Assink & Wibbelink, 2016). Besides, existing effect size transformation methods were applied to homogenize the effect sizes and enhance the interpretability. Finally, we tested the robustness and explored the heterogeneity through sensitivity and moderator meta-analysis.

Nevertheless, several limitations should be noted. Substantial heterogeneity between studies persisted in the meta-analysis of pesticide exposure and telomere length, despite the implementation of moderator analysis, and this might have prevented producing reliable results. Secondly, the predominance of cross-sectional studies included weakened the pooled level of evidence. Thirdly, only two studies were included in the meta-analysis of the epigenetic clock, allowing for effect estimates being reported as continuous beta coefficients on the same epigenetic clock. Fourthly, a large portion of the studies only reported broad categories of pesticide mixtures rather than the active chemical ingredients, making it difficult to evaluate individual substances. Lastly, only two biomarkers of biological aging were included in this systematic review, restricting our ability to generalize the results to the overall aging process.

## 5. Conclusion

This systematic review and meta-analysis do not allow a final conclusion on the association between pesticide exposure and biological ageing measured as epigenetic clock or as telomere length. However, both analyses are suggestive of a possible positive association between pesticide exposure and accelerated aging, although this is not always firmly supported by statistically significant pooled effect estimates. Inadequate study quality, high heterogeneity of reported results, publication bias in studies reporting on telomere length, and the small number of studies reporting on epigenetic clocks, are the main constraints limiting our inference. Future studies conducted on large samples coming from high-quality cohort studies, integrate more biomarkers of biological aging and assessing exposure to pesticides through biomarkers with refined adjustments for potential confounders are warranted.

## Supporting information

file:///C:/Users/P310419/Desktop/Manuscript/Preprint/Supplementary%20Material.htm

## Data Availability

All data produced in the present study are available upon reasonable request to the authors

## CRediT authorship contribution statement

**Shanshan Zuo**: Methodology, Software, Formal analysis, Investigation, Writing - Original Draft, Visualization; **Vidhya Sasitharan**: Investigation, Writing - Review & Editing; **Gian Luca Di Tanna**: Writing - Review & Editing; **Judith M. Vonk**: Methodology, Writing - Review & Editing, Supervision; **Maaike De Vries**: Writing - Review & Editing; **Moustafa Sherif**: Writing - Review & Editing; **Balázs Ádám**: Writing - Review & Editing; **Juan Carlos Rivillas**: Writing - Review & Editing; **Valentina Gallo**: Conceptualization, Investigation, Writing - Review & Editing, Supervision.

## Data availability

Data will be made available on request.

## Acknowledgements

Shanshan Zuo is supported by China Scholarship Council (202208620033). The authors would like to thank Lauren B. Ellis at Northeastern University for providing suggestions on data transformation and synthesis. Similarly, the authors would like to thank the appointed librarian at the University of Groningen, Joost Driesens, who provided guidance on the search terms selection and search engines. Furthermore, the authors appreciate the authors who supplied the necessary data.

