## Supplementary material for "Is exposure to pesticides associated with biological aging? A systematic review and meta-analysis": file:///C:/Users/P310419/Desktop/Manuscript/Preprint/Supplementary%20Material.htm

Appendix A

Table A.1 PRISMA checklist

Table A.2 Search strategy

Table A.3 Results of included studies on pesticide exposure and epigenetic clocks.

Table A.4 Results of included studies on pesticide exposure and telomere length.

Table A.5 Risk of bias assessment by adapted Newcastle–Ottawa Scale for cross-sectional studies.

Table A.6 Risk of bias assessment by adapted Newcastle–Ottawa Quality Assessment Scale for cohort studies

Fig. A.1 Funnel plot of associations between pesticide exposure and telomere length

Appendix B

Formulas for effect estimate calculation and transformation

Appendix A

Table A. 1 PRISMA checklist

| **Section and Topic** | **Item #** | **Checklist item** | **Location where item is reported** |
| --- | --- | --- | --- |
| **TITLE** | | |  |
| Title | 1 | Identify the report as a systematic review. | P1 |
| **ABSTRACT** | | |  |
| Abstract | 2 | See the PRISMA 2020 for Abstracts checklist. | P2 |
| **INTRODUCTION** | | |  |
| Rationale | 3 | Describe the rationale for the review in the context of existing knowledge. | P3-4 |
| Objectives | 4 | Provide an explicit statement of the objective(s) or question(s) the review addresses. | P4 |
| **METHODS** | | |  |
| Eligibility criteria | 5 | Specify the inclusion and exclusion criteria for the review and how studies were grouped for the syntheses. | P4, P22 |
| Information sources | 6 | Specify all databases, registers, websites, organisations, reference lists and other sources searched or consulted to identify studies. Specify the date when each source was last searched or consulted. | P4 |
| Search strategy | 7 | Present the full search strategies for all databases, registers and websites, including any filters and limits used. | P4, Table A.2 |
| Selection process | 8 | Specify the methods used to decide whether a study met the inclusion criteria of the review, including how many reviewers screened each record and each report retrieved, whether they worked independently, and if applicable, details of automation tools used in the process. | P4-5 |
| Data collection process | 9 | Specify the methods used to collect data from reports, including how many reviewers collected data from each report, whether they worked independently, any processes for obtaining or confirming data from study investigators, and if applicable, details of automation tools used in the process. | P4-5 |
| Data items | 10a | List and define all outcomes for which data were sought. Specify whether all results that were compatible with each outcome domain in each study were sought (e.g. for all measures, time points, analyses), and if not, the methods used to decide which results to collect. | P4-5 |
|  | 10b | List and define all other variables for which data were sought (e.g. participant and intervention characteristics, funding sources). Describe any assumptions made about any missing or unclear information. | P4-5 |
| Study risk of bias assessment | 11 | Specify the methods used to assess risk of bias in the included studies, including details of the tool(s) used, how many reviewers assessed each study and whether they worked independently, and if applicable, details of automation tools used in the process. | P5 |
| Effect measures | 12 | Specify for each outcome the effect measure(s) (e.g. risk ratio, mean difference) used in the synthesis or presentation of results. | P5 |
| Synthesis methods | 13a | Describe the processes used to decide which studies were eligible for each synthesis (e.g. tabulating the study intervention characteristics and comparing against the planned groups for each synthesis (item #5)). | P5-6 |
|  | 13b | Describe any methods required to prepare the data for presentation or synthesis, such as handling of missing summary statistics, or data conversions. | Appendix B |
|  | 13c | Describe any methods used to tabulate or visually display results of individual studies and syntheses. | P5-6 |
|  | 13d | Describe any methods used to synthesize results and provide a rationale for the choice(s). If meta-analysis was performed, describe the model(s), method(s) to identify the presence and extent of statistical heterogeneity, and software package(s) used. | P5-6 |
|  | 13e | Describe any methods used to explore possible causes of heterogeneity among study results (e.g. subgroup analysis, meta-regression). | P6 |
|  | 13f | Describe any sensitivity analyses conducted to assess robustness of the synthesized results. | P6 |
| Reporting bias assessment | 14 | Describe any methods used to assess risk of bias due to missing results in a synthesis (arising from reporting biases). | P6 |
| Certainty assessment | 15 | Describe any methods used to assess certainty (or confidence) in the body of evidence for an outcome. | NA |
| **RESULTS** | | |  |
| Study selection | 16a | Describe the results of the search and selection process, from the number of records identified in the search to the number of studies included in the review, ideally using a flow diagram. | Fig. 1 |
|  | 16b | Cite studies that might appear to meet the inclusion criteria, but which were excluded, and explain why they were excluded. | Fig. 1 |
| Study characteristics | 17 | Cite each included study and present its characteristics. | Table 2-3 |
| Risk of bias in studies | 18 | Present assessments of risk of bias for each included study. | Table A.5, Table A.6 |
| Results of individual studies | 19 | For all outcomes, present, for each study: (a) summary statistics for each group (where appropriate) and (b) an effect estimate and its precision (e.g. confidence/credible interval), ideally using structured tables or plots. | Table A. 2, Table A.4 |
| Results of syntheses | 20a | For each synthesis, briefly summarise the characteristics and risk of bias among contributing studies. | P6-7 |
|  | 20b | Present results of all statistical syntheses conducted. If meta-analysis was done, present for each the summary estimate and its precision (e.g. confidence/credible interval) and measures of statistical heterogeneity. If comparing groups, describe the direction of the effect. | Fig. 2, Fig. 3 |
|  | 20c | Present results of all investigations of possible causes of heterogeneity among study results. | Table 5 |
|  | 20d | Present results of all sensitivity analyses conducted to assess the robustness of the synthesized results. | Table 4 |
| Reporting biases | 21 | Present assessments of risk of bias due to missing results (arising from reporting biases) for each synthesis assessed. | Fig. A.1 |
| Certainty of evidence | 22 | Present assessments of certainty (or confidence) in the body of evidence for each outcome assessed. | NA |
| **DISCUSSION** | | |  |
| Discussion | 23a | Provide a general interpretation of the results in the context of other evidence. | P10-12 |
|  | 23b | Discuss any limitations of the evidence included in the review. | P11-13 |
|  | 23c | Discuss any limitations of the review processes used. | P13 |
|  | 23d | Discuss implications of the results for practice, policy, and future research. | P13 |
| **OTHER INFORMATION** | | |  |
| Registration and protocol | 24a | Provide registration information for the review, including register name and registration number, or state that the review was not registered. | P4 |
|  | 24b | Indicate where the review protocol can be accessed, or state that a protocol was not prepared. | P4 |
|  | 24c | Describe and explain any amendments to information provided at registration or in the protocol. | NA |
| Support | 25 | Describe sources of financial or non-financial support for the review, and the role of the funders or sponsors in the review. | P14 |
| Competing interests | 26 | Declare any competing interests of review authors. | P14 |
| Availability of data, code and other materials | 27 | Report which of the following are publicly available and where they can be found: template data collection forms; data extracted from included studies; data used for all analyses; analytic code; any other materials used in the review. | P14 |

Table A. 2 Search strategy

**Pubmed**

("Pesticides"[MeSH Terms] OR ("pesticides"[Title/Abstract] OR "insecticides"[Title/Abstract] OR "herbicides"[Title/Abstract] OR "fungicides"[Title/Abstract] OR "bactericides"[Title/Abstract] OR "rodenticides"[Title/Abstract] OR "fumigants"[Title/Abstract])) AND ("biological ag*"[Title/Abstract] OR "senescence"[Title/Abstract] OR "epigenetic clock"[Title/Abstract] OR "epigenetic ag*"[Title/Abstract] OR "methylation ag*"[Title/Abstract] OR "epigenetic age acceleration"[Title/Abstract] OR "telomere"[Title/Abstract] OR "telomere length"[Title/Abstract] OR "klemera doubal"[Title/Abstract] OR "homeostatic dysregulation"[Title/Abstract] OR "allostatic load"[Title/Abstract] OR "biological health score"[Title/Abstract] OR ("methyl*"[Title/Abstract] AND ("Horvath*"[Title/Abstract] OR "Hannum*"[Title/Abstract] OR "PhenoAge*"[Title/Abstract] OR "GrimAge*"[Title/Abstract] OR "Dunedin*"[Title/Abstract])))

**Embase**

(('pesticide'/exp OR pesticide OR insecticide:ti,ab,kw OR herbicide:ti,ab,kw OR fungicide:ti,ab,kw OR bactericide:ti,ab,kw OR rodenticide:ti,ab,kw OR fumigant:ti,ab,kw)) AND (('biological ag*':ti,ab,kw OR senescence:ti,ab,kw OR 'epigenetic clock':ti,ab,kw OR 'epigenetic ag*':ti,ab,kw OR 'methylation ag*':ti,ab,kw OR 'epigenetic age acceleration':ti,ab,kw OR telomere:ti,ab,kw OR 'telomere length':ti,ab,kw OR 'klemera doubal':ti,ab,kw OR 'homeostatic dysregulation':ti,ab,kw OR 'allostatic load':ti,ab,kw OR 'biological health score':ti,ab,kw) OR (methyl*:ti,ab,kw AND (horvath*:ti,ab,kw OR hannum*:ti,ab,kw OR phenoage*:ti,ab,kw OR grimage*:ti,ab,kw OR dunedin*:ti,ab,kw)))

**Web of Science**

[((((((TS=(pesticides)) OR TS=(insecticides)) OR TS=(herbicides)) OR TS=(fungicides)) OR TS=(bactericides)) OR TS=(rodenticides)) OR TS=(fumigants)] AND [[(((((((((((TS=("biological ag*")) OR TS=(senescence)) OR TS=("epigenetic clock")) OR TS=("epigenetic ag*")) OR TS=("methylation ag*")) OR TS=("epigenetic age acceleration")) OR TS=(telomere)) OR TS=("telomere length")) OR TS=("klemera doubal"))] OR [TS=("homeostatic dysregulation")) OR TS=("allostatic load")) OR TS=("biological health score") OR [TS=(methyl*) AND ((((TS=(Horvath*)) OR TS=(Hannum*)) OR TS=(PhenoAge*)) OR TS=(GrimAge*)) OR TS=(Dunedin*)]]]

Table A. 3 Results of included studies on pesticide exposure and epigenetic clocks.

| Author, year | Pesticides | Clocks | β | LL | UL | P value | Associations |
| --- | --- | --- | --- | --- | --- | --- | --- |
| Lucia, et al., 2022 | Glyphosate | Horvath | -0.33 | -0.75 | 0.10 | 0.13 | AMPA was associated with increased epigenetic age acceleration according to the Hannum and Horvath clocks, while glyphosate was not significantly associated with any of the epigenetic clocks. |
|  |  | Hannum | -0.22 | -0.58 | 0.15 | 0.24 |  |
|  |  | PhenoAge | 0.03 | -0.60 | 0.66 | 0.93 |  |
|  |  | DunedinPoAm | 0.07 | -0.02 | 0.16 | 0.11 |  |
|  | AMPA | Horvath | **0.41** | **0.02** | **0.80** | 0.04 |  |
|  |  | Hannum | **0.36** | **0.03** | **0.7** | 0.04 |  |
|  |  | PhenoAge | 0.3 | -0.29 | 0.88 | 0.32 |  |
|  |  | DunedinPoAm | 0.01 | -0.07 | 0.09 | 0.85 |  |
| Lind, et al., 2018 | p,p′-DDE | Hannum | **0.44** | **0.13** | **0.75** | 0.0051 | Increased p,p′-DDE and TNC, but not HCB, were related to increased epigenetic age |
|  | HCB |  | 0.18 | -0.51 | 0.87 | 0.60 |  |
|  | TNC |  | **0.66** | **0.15** | **1.17** | 0.011 |  |
| De Prado-Bert, et al., 2021 | DDE | Horvath’s Skin and Blood | -0.02 | -0.09 | 0.04 | 0.484 | Exposure to the DMDTP was associated with decreased age acceleration |
|  | DDT |  | -0.01 | -0.08 | 0.06 | 0.741 |  |
|  | HCB |  | -0.06 | -0.12 | 0.003 | 0.062 |  |
|  | DEP |  | -0.03 | -0.1 | 0.03 | 0.268 |  |
|  | DETP |  | -0.07 | -0.14 | 0.00 | 0.052 |  |
|  | DMP |  | 0.03 | -0.04 | 0.10 | 0.365 |  |
|  | DMTP |  | 0.00 | -0.05 | 0.06 | 0.882 |  |
|  | DMDTP |  | **-0.13** | **-0.24** | **-0.02** | **0.017** |  |
| Hoang, et al., 2021 | Dicamba | Horvath | 0.66 | -1.14 | 2.46 | 0.47 | Past use of DDT was associated with positive age acceleration based on the Hannum and Skin and Blood clocks |
|  |  | Hannum | 0.34 | -1.40 | 2.08 | 0.70 |  |
|  |  | PhenoAge | 0.24 | -2.03 | 2.51 | 0.83 |  |
|  |  | Skin and Blood | 0.24 | -1.40 | 1.88 | 0.77 |  |
|  |  | GrimAge | -0.18 | -1.18 | 0.82 | 0.72 |  |
|  | Picloram | Horvath | 0.86 | -1.11 | 2.83 | 0.39 |  |
|  |  | Hannum | -0.12 | -2.09 | 1.85 | 0.91 |  |
|  |  | PhenoAge | -1.62 | -4.12 | 0.87 | 0.20 |  |
|  |  | Skin and Blood | 0.66 | -1.13 | 2.45 | 0.47 |  |
|  |  | GrimAge | 0.00 | -1.09 | 1.08 | 0.10 |  |
|  | Mesotrione | Horvath | 0.11 | -1.74 | 1.96 | 0.91 |  |
|  |  | Hannum | -0.77 | -2.60 | 1.07 | 0.41 |  |
|  |  | PhenoAge | 0.07 | -2.26 | 2.39 | 0.95 |  |
|  |  | Skin and Blood | -0.25 | -1.93 | 1.42 | 0.77 |  |
|  |  | GrimAge | 0.18 | -0.83 | 1.19 | 0.73 |  |
|  | Acetochlor | Horvath | 0.16 | -1.62 | 1.94 | 0.86 |  |
|  |  | Hannum | 0.82 | -0.96 | 2.60 | 0.37 |  |
|  |  | PhenoAge | -0.01 | -2.26 | 2.25 | 1.00 |  |
|  |  | Skin and Blood | 0.32 | -1.32 | 1.96 | 0.70 |  |
|  |  | GrimAge | -0.37 | -1.36 | 0.62 | 0.47 |  |
|  | Metolachlor | Horvath | -0.38 | -2.07 | 1.30 | 0.66 |  |
|  |  | Hannum | -0.26 | -1.83 | 1.30 | 0.74 |  |
|  |  | PhenoAge | -0.10 | -2.09 | 1.90 | 0.92 |  |
|  |  | Skin and Blood | -1.10 | -2.57 | 0.37 | 0.14 |  |
|  |  | GrimAge | -0.03 | -0.95 | 0.90 | 0.95 |  |
|  | Glyphosate | Horvath | 0.37 | -1.16 | 1.89 | 0.64 |  |
|  |  | Hannum | -0.08 | -1.60 | 1.44 | 0.92 |  |
|  |  | PhenoAge | -1.21 | -3.09 | 0.68 | 0.21 |  |
|  |  | Skin and Blood | -0.21 | -1.56 | 1.14 | 0.76 |  |
|  |  | GrimAge | 0.07 | -0.72 | 0.86 | 0.86 |  |
|  | 2.4-D | Horvath | 0.16 | -0.84 | 1.17 | 0.75 |  |
|  |  | Hannum | 0.15 | -0.84 | 1.14 | 0.76 |  |
|  |  | PhenoAge | 0.86 | -0.40 | 2.12 | 0.18 |  |
|  |  | Skin and Blood | 0.32 | -0.58 | 1.23 | 0.49 |  |
|  |  | GrimAge | 0.02 | -0.52 | 0.57 | 0.93 |  |
|  | Atrazine | Horvath | 0.64 | -0.82 | 2.10 | 0.39 |  |
|  |  | Hannum | 0.69 | -0.74 | 2.13 | 0.34 |  |
|  |  | PhenoAge | 1.08 | -0.71 | 2.87 | 0.24 |  |
|  |  | Skin and Blood | 1.11 | -0.14 | 2.37 | 0.08 |  |
|  |  | GrimAge | 0.25 | -0.54 | 1.05 | 0.53 |  |
|  | Malathion | Horvath | -1.18 | -3.20 | 0.84 | 0.25 |  |
|  |  | Hannum | -0.51 | -2.36 | 1.34 | 0.59 |  |
|  |  | PhenoAge | -0.43 | -2.91 | 2.04 | 0.73 |  |
|  |  | Skin and Blood | 0.55 | -1.27 | 2.36 | 0.56 |  |
|  |  | GrimAge | -0.29 | -1.30 | 0.73 | 0.58 |  |
|  | Aldrin | Horvath | 0.51 | -0.31 | 1.32 | 0.22 |  |
|  |  | Hannum | -0.15 | -0.96 | 0.65 | 0.71 |  |
|  |  | PhenoAge | -0.32 | -1.34 | 0.70 | 0.54 |  |
|  |  | Skin and Blood | -0.15 | -0.89 | 0.58 | 0.68 |  |
|  |  | GrimAge | -0.20 | -0.65 | 0.24 | 0.37 |  |
|  | Chlordane | Horvath | -0.20 | -0.94 | 0.53 | 0.58 |  |
|  |  | Hannum | -0.04 | -0.76 | 0.69 | 0.92 |  |
|  |  | PhenoAge | -0.69 | -1.61 | 0.22 | 0.14 |  |
|  |  | Skin and Blood | 0.21 | -0.45 | 0.87 | 0.53 |  |
|  |  | GrimAge | **-0.46** | **-0.86** | **-0.07** | 0.02 |  |
|  | DDT | Horvath | 0.44 | -0.38 | 1.26 | 0.30 |  |
|  |  | Hannum | **1.15** | **0.34** | **1.96** | 0.01 |  |
|  |  | PhenoAge | 0.77 | -0.26 | 1.80 | 0.14 |  |
|  |  | Skin and Blood | **0.81** | **0.07** | **1.55** | 0.03 |  |
|  |  | GrimAge | 0.19 | -0.26 | 0.63 | 0.41 |  |
|  | Dieldrin | Horvath | 0.57 | -0.59 | 1.73 | 0.33 |  |
|  |  | Hannum | 0.89 | -0.26 | 2.03 | 0.13 |  |
|  |  | PhenoAge | -0.06 | -1.52 | 1.39 | 0.93 |  |
|  |  | Skin and Blood | 0.45 | -0.59 | 1.50 | 0.40 |  |
|  |  | GrimAge | 0.13 | -0.50 | 0.76 | 0.69 |  |
|  | Heptachlor | Horvath | 0.73 | -0.14 | 1.60 | 0.10 |  |
|  |  | Hannum | 0.66 | -0.20 | 1.52 | 0.13 |  |
|  |  | PhenoAge | 0.32 | -0.77 | 1.42 | 0.56 |  |
|  |  | Skin and Blood | 0.61 | -0.18 | 1.39 | 0.13 |  |
|  |  | GrimAge | -0.14 | -0.62 | 0.33 | 0.55 |  |
|  | Lindane | Horvath | -0.33 | -1.06 | 0.40 | 0.38 |  |
|  |  | Hannum | -0.43 | -1.15 | 0.30 | 0.25 |  |
|  |  | PhenoAge | -0.41 | -1.33 | 0.50 | 0.38 |  |
|  |  | Skin and Blood | 0.03 | -0.63 | 0.70 | 0.92 |  |
|  |  | GrimAge | -0.37 | -0.77 | 0.03 | 0.07 |  |
|  | Toxaphene | Horvath | -0.37 | -1.24 | 0.50 | 0.41 |  |
|  |  | Hannum | 0.21 | -0.64 | 1.07 | 0.62 |  |
|  |  | PhenoAge | -0.10 | -1.18 | 0.99 | 0.86 |  |
|  |  | Skin and Blood | 0.13 | -0.65 | 0.91 | 0.74 |  |
|  |  | GrimAge | -0.17 | -0.65 | 0.30 | 0.47 |  |

Abbreviations: HCB: hexachlorobenzene; TNC: transnonachlor; DMP: dimethylphosphate; DEP: diethylphosphate; DMTP: dimethylthiophosphate; DETP: diethylthiophosphate; DMDTP: dimethyldithiophosphate; DEDTP: diethyldithiophosphate; TCPY: 3.5.6-trichloro-2-pyridinol; LL: lower limit; UL: upper limit. Bold indicates a statistically significant estimate.

Table A. 4 Results of included studies on pesticide exposure and telomere length.

| Author, year | Pesticides | NE | ME | SE | NN | MN | SN | β (raw)/r | β (percent change)/r | LL | UL | P value | Associations |
| --- | --- | --- | --- | --- | --- | --- | --- | --- | --- | --- | --- | --- | --- |
| Dos Santos, et al., 2022 | Mixture including herbicides, insecticides and fungicides | 81 | 50.84 | 31.95 | 81 | 39.65 | 16.67 |  |  |  |  |  | No difference was observed for TL between groups |
| Duan, et al., 2017 | Omethoate | 180 | 1.73 | 1.09 | 115 | 1.01 | 0.42 |  |  |  |  |  | Relative telomere lengths in the exposure group were significantly longer than that in the control group |
| Saad-Hussein, et al., 2019 | Mixture compounds, mostly Ops (malathionchloropyrifos and dimethoate) and carbamates (carbofuran) | 100 | 0.68 | 0.04 | 100 | 0.84 | 0.03 |  |  |  |  |  | Shortening of telomere length was found in pesticides-exposed subjects compared to their controls |
| De Oliveira, et al., 2019 | Mixture including fungicide, herbicide, insecticide | 76 | 8688 | 3628 | 72 | 7463 | 3409 |  |  |  |  |  | No difference in the telomere length in the exposed group compared to the non-exposed group. |
| Kahl, et al., 2018a | Mixture including carbamates, organophosphates, pyrethroids and organochlorines | 121 | 3937 | 635 | 121 | 6387 | 3127 |  |  |  |  |  | Reduced telomere length was found in farmers compared to non-exposed individuals |
| Kahl, et al., 2018b | Mixture including fungicide, herbicide, insecticide | 40 | 4098 | 666 | 40 | 4551 | 920 |  |  |  |  |  | Telomere length measured was shorter in exposed individuals |
| Kahl, et al., 2018c | Mixture | 56 | 4615 | 1189 | 74 | 5725 | 1404 |  |  |  |  |  | Exposed group showed significantly shorter telomeres |
| Kahl, et al., 2016 | Mixture including glyphosate and flumetralin | 62 | 32.10 | 12.60 | 62 | 47.00 | 19.80 |  |  |  |  |  | Significant decrease in telomere length was found in the exposed relative to the non-exposed group |
| Ali. et al., 2023 | DMP | 71 | 1.22 | 1.01 | 71 | 1.22 | 0.86 |  |  |  |  |  | Women with high urine levels of DEP. DETP or DEDTP and living in proximity to agricultural fields had shorter telomeres |
|  | DEP | 71 | 1.07 | 0.79 | 71 | 1.37 | 1.04 |  |  |  |  |  |  |
|  | DMTP | 71 | 1.21 | 0.93 | 71 | 1.23 | 0.91 |  |  |  |  |  |  |
|  | DETP | 71 | 0.98 | 0.76 | 71 | 1.46 | 1.07 |  |  |  |  |  |  |
|  | DMDTP | 71 | 1.2 | 0.91 | 71 | 1.24 | 0.93 |  |  |  |  |  |  |
|  | DEDTP | 71 | 0.92 | 0.66 | 71 | 1.52 | 1.07 |  |  |  |  |  |  |
| Hou. et al., 2013 | Alachlor | 659 | 1.17 | 0.32 | 466 | 1.24 | 0.41 |  |  |  |  |  | Seven pesticides (alachlor. metolachlor. tri­ fluralin. 2.4­D. permethrin. toxaphene and DDT) were negatively associated with telomere length among pesticide applicators |
|  | Metolachlor | 492 | 1.17 | 0.33 | 622 | 1.23 | 0.38 |  |  |  |  |  |  |
|  | Trifluralin | 599 | 1.18 | 0.36 | 515 | 1.23 | 0.36 |  |  |  |  |  |  |
|  | 2.4-D | 1004 | 1.19 | 0.33 | 194 | 1.27 | 0.48 |  |  |  |  |  |  |
|  | DDT | 371 | 1.14 | 0.32 | 428 | 1.21 | 0.34 |  |  |  |  |  |  |
|  | Permethrin (poultry/livestock) | 104 | 1.15 | 0.27 | 1021 | 1.21 | 0.37 |  |  |  |  |  |  |
|  | Toxaphene | 128 | 1.14 | 0.33 | 679 | 1.18 | 0.33 |  |  |  |  |  |  |
|  | Atrazine | 884 | 1.20 | 0.36 | 320 | 1.21 | 0.35 |  |  |  |  |  |  |
|  | Butylate | 218 | 1.19 | 0.33 | 592 | 1.18 | 0.34 |  |  |  |  |  |  |
|  | Chlorimuron-ethyl | 225 | 1.16 | 0.31 | 586 | 1.19 | 0.35 |  |  |  |  |  |  |
|  | Cyanazine | 509 | 1.19 | 0.33 | 610 | 1.22 | 0.39 |  |  |  |  |  |  |
|  | Dicamba | 602 | 1.17 | 0.32 | 510 | 1.23 | 0.42 |  |  |  |  |  |  |
|  | EPTC | 207 | 1.17 | 0.31 | 899 | 1.21 | 0.37 |  |  |  |  |  |  |
|  | Glyphosate | 920 | 1.20 | 0.37 | 291 | 1.19 | 0.33 |  |  |  |  |  |  |
|  | Imazethapyr | 435 | 1.20 | 0.31 | 669 | 1.2 | 0.38 |  |  |  |  |  |  |
|  | Metribuzin | 319 | 1.16 | 0.31 | 490 | 1.19 | 0.35 |  |  |  |  |  |  |
|  | Paraquat | 144 | 1.21 | 0.35 | 666 | 1.17 | 0.33 |  |  |  |  |  |  |
|  | Pendimethalin | 270 | 1.18 | 0.34 | 548 | 1.18 | 0.33 |  |  |  |  |  |  |
|  | Petroleum oils | 185 | 1.19 | 0.30 | 612 | 1.18 | 0.34 |  |  |  |  |  |  |
|  | 2.4.5 trichlorophenoxy acetic acid | 267 | 1.14 | 0.31 | 538 | 1.19 | 0.34 |  |  |  |  |  |  |
|  | 2.4.5 TP | 72 | 1.16 | 0.28 | 734 | 1.18 | 0.3 |  |  |  |  |  |  |
|  | Aldicarb | 42 | 1.22 | 0.35 | 770 | 1.18 | 0.33 |  |  |  |  |  |  |
|  | Aldrin | 262 | 1.15 | 0.30 | 538 | 1.19 | 0.35 |  |  |  |  |  |  |
|  | Carbaryl | 387 | 1.21 | 0.34 | 417 | 1.15 | 0.3 |  |  |  |  |  |  |
|  | Carbofuran | 387 | 1.20 | 0.33 | 726 | 1.21 | 0.38 |  |  |  |  |  |  |
|  | Chlordane | 245 | 1.16 | 0.34 | 556 | 1.18 | 0.33 |  |  |  |  |  |  |
|  | Chlorpyrifos | 473 | 1.21 | 0.38 | 736 | 1.19 | 0.35 |  |  |  |  |  |  |
|  | Coumaphos | 114 | 1.21 | 0.36 | 974 | 1.2 | 0.36 |  |  |  |  |  |  |
|  | Diazinon | 192 | 1.21 | 0.32 | 608 | 1.17 | 0.33 |  |  |  |  |  |  |
|  | Dichlorvos | 167 | 1.18 | 0.33 | 947 | 1.21 | 0.36 |  |  |  |  |  |  |
|  | Dieldrin | 63 | 1.14 | 0.29 | 742 | 1.18 | 0.33 |  |  |  |  |  |  |
|  | Fonofos | 249 | 1.19 | 0.34 | 870 | 1.21 | 0.37 |  |  |  |  |  |  |
|  | Heptachlor | 201 | 1.15 | 0.30 | 608 | 1.19 | 0.35 |  |  |  |  |  |  |
|  | Lindane | 139 | 1.16 | 0.30 | 663 | 1.18 | 0.34 |  |  |  |  |  |  |
|  | Malathion | 535 | 1.18 | 0.35 | 265 | 1.17 | 0.3 |  |  |  |  |  |  |
|  | Parathion | 79 | 1.20 | 0.32 | 718 | 1.17 | 0.32 |  |  |  |  |  |  |
|  | Permethrin (for crop) | 109 | 1.22 | 0.34 | 993 | 1.2 | 0.37 |  |  |  |  |  |  |
|  | Phorate | 273 | 1.15 | 0.30 | 537 | 1.19 | 0.34 |  |  |  |  |  |  |
|  | Terbufos | 423 | 1.20 | 0.32 | 695 | 1.2 | 0.39 |  |  |  |  |  |  |
|  | Aluminum phosphide | 32 | 1.16 | 0.18 | 780 | 1.17 | 0.33 |  |  |  |  |  |  |
|  | Methyl bromide | 185 | 1.23 | 0.47 | 1034 | 1.2 | 0.33 |  |  |  |  |  |  |
|  | Ethylene dibromide | 40 | 1.28 | 0.60 | 767 | 1.17 | 0.31 |  |  |  |  |  |  |
|  | Carbon tetrachloride/Carbon disulfide–80/20 mix | 73 | 1.15 | 0.38 | 734 | 1.18 | 0.33 |  |  |  |  |  |  |
|  | Benomyl | 59 | 1.24 | 0.33 | 743 | 1.17 | 0.33 |  |  |  |  |  |  |
|  | Captan | 116 | 1.16 | 0.32 | 968 | 1.21 | 0.36 |  |  |  |  |  |  |
|  | Chlorothalonil | 81 | 1.21 | 0.37 | 1132 | 1.2 | 0.35 |  |  |  |  |  |  |
|  | Maneb/Mancozeb | 61 | 1.26 | 0.47 | 737 | 1.17 | 0.32 |  |  |  |  |  |  |
|  | Metalaxyl | 120 | 1.22 | 0.41 | 675 | 1.17 | 0.31 |  |  |  |  |  |  |
| Andreotti et al., 2015 | Alachlor | 381 | 1.11 | 0.59 | 187 | 1.04 | 0.55 |  |  |  |  |  | Increasing lifetime days of 2.4-D. diazinon. and butylate were significantly associated with shorter telomere length while alachlor was significantly associated with longer telomere length |
|  | Butylate | 224 | 1.02 | 0.60 | 344 | 1.1 | 0.56 |  |  |  |  |  |  |
|  | Metolachlor | 301 | 1.11 | 0.69 | 267 | 1.04 | 0.49 |  |  |  |  |  |  |
|  | Paraquat | 207 | 1.1 | 0.58 | 361 | 1.12 | 0.76 |  |  |  |  |  |  |
|  | Sethoxydim | 38 | 1.23 | 0.55 | 530 | 1.1 | 0.69 |  |  |  |  |  |  |
|  | 2.4-D | 469 | 1.03 | 0.65 | 99 | 1.22 | 0.60 |  |  |  |  |  |  |
|  | 2.4.5-TP | 84 | 1.25 | 0.55 | 484 | 1.08 | 0.66 |  |  |  |  |  |  |
|  | Aldrin | 139 | 1.04 | 0.59 | 429 | 1.13 | 0.83 |  |  |  |  |  |  |
|  | DDT | 213 | 1.17 | 0.58 | 355 | 0.99 | 0.75 |  |  |  |  |  |  |
|  | Diazinon | 284 | 1.06 | 0.51 | 284 | 1.15 | 0.67 |  |  |  |  |  |  |
|  | Heptachlor | 114 | 1.11 | 0.53 | 454 | 1.05 | 0.64 |  |  |  |  |  |  |
|  | Benomyl | 137 | 1.15 | 0.59 | 431 | 1.13 | 1.04 |  |  |  |  |  |  |
|  | Chlorothalonil | 104 | 1.14 | 0.61 | 464 | 1.1 | 0.65 |  |  |  |  |  |  |
|  | Maneb/Mancozeb | 91 | 1.19 | 0.57 | 477 | 1.08 | 0.66 |  |  |  |  |  |  |
|  | Metalaxyl | 194 | 1.13 | 0.56 | 374 | 1.09 | 0.77 |  |  |  |  |  |  |
| Cosemans. et al., 2022 | Glyphosate |  |  |  |  |  |  |  | 3.31 | -1.17 | 8.07 | 0.15 | AMPA concentration was associated with longer telomere length. while no association was observed with glyphosate |
|  | AMPA |  |  |  |  |  |  |  | 5.19 | 0.49 | 10.11 | 0.03 |  |
| Guzzardi. et al., 2016 | Oxychlordane |  |  |  |  |  |  | -0.062 |  |  |  | 0.052 | Exposure to oxychlordane and trans-nonachlor predicts telomere attrition |
|  | Trans-nonachlor |  |  |  |  |  |  | -0.068 |  |  |  | 0.030 |  |
|  | p. p'-DDE |  |  |  |  |  |  | -0.03 |  |  |  | 0.325 |  |
| Karimi. et al., 2020 | HCB |  |  |  |  |  |  |  | -4.75 | -8.44 | -1.19 |  | HCB, Heptachlor epoxide, Heptachlor, Aldrin, DDD and DDE were negatively associated with telomere length |
|  | Heptachlor epoxide |  |  |  |  |  |  |  | -7.62 | -11.69 | -4.38 |  |  |
|  | Heptachlor |  |  |  |  |  |  |  | -11.56 | -15.44 | -7.62 |  |  |
|  | Aldrin |  |  |  |  |  |  |  | -10.63 | -14.44 | -7.19 |  |  |
|  | DDT |  |  |  |  |  |  |  | -1.00 | -2.19 | 0.03 |  |  |
|  | DDD |  |  |  |  |  |  |  | -4.94 | -6.81 | -3.22 |  |  |
|  | DDE |  |  |  |  |  |  |  | -3.37 | -5.00 | -1.69 |  |  |
| Ock. et al., 2020 | DEP |  |  |  |  |  |  | 0.03 |  | -0.01 | 0.07 |  | Shorter telomere lengths were associated with TCPY and longer telomere lengths were associated with DETP. |
|  | DMTP |  |  |  |  |  |  | -0.01 |  | -0.05 | 0.03 |  |  |
|  | DMP |  |  |  |  |  |  | 0.00 |  | -0.03 | 0.04 |  |  |
|  | DMDTP |  |  |  |  |  |  | 0.00 |  | -0.03 | 0.04 |  |  |
|  | DEDTP |  |  |  |  |  |  | 0.02 |  | -0.04 | 0.07 |  |  |
|  | TCPY |  |  |  |  |  |  | 0.02 |  | -0.03 | 0.08 |  |  |
|  | DETP |  |  |  |  |  |  | 0.06 |  | 0.01 | 0.11 |  |  |
| Shin. et al., 2010 | p.p'-DDE |  |  |  |  |  |  | 0.31 |  |  |  |  | Low-dose exposure to organochlorine pesticides was associated with increased telomere length in healthy populations. However, within POPs concentrations over certain levels. POPs were associated with decreased telomere length |
|  | p.p'-DDT |  |  |  |  |  |  | 0.15 |  |  |  |  |  |
|  | p.p'-DDD |  |  |  |  |  |  | -0.09 |  |  |  |  |  |
|  | oxychlordane |  |  |  |  |  |  | 0.24 |  |  |  |  |  |
|  | trans-nonachlor |  |  |  |  |  |  | 0.23 |  |  |  |  |  |
|  | heptachlor epoxide |  |  |  |  |  |  | 0.27 |  |  |  |  |  |
|  | β-Hexachlorocyclohexane |  |  |  |  |  |  | 0.16 |  |  |  |  |  |
|  | Hexachlorobenzene |  |  |  |  |  |  | 0.13 |  |  |  |  |  |
|  | Mirex |  |  |  |  |  |  | 0.10 |  |  |  |  |  |

Abbreviations: NE: number of exposed group; NN: number of non-exposed group; OPs: organophosphates; ME: mean telomere length of exposed group; MN: mean telomere length of non-exposed group; SE: standard deviation of exposed group; SN: standard deviation of non-exposed group; HCB: hexachlorobenzene; TNC: transnonachlor; DMP: dimethylphosphate; DEP: diethylphosphate; DMTP: dimethylthiophosphate; DETP: diethylthiophosphate; DMDTP: dimethyldithiophosphate; DEDTP: diethyldithiophosphate; TCPY: 3.5.6-trichloro-2-pyridinol; LL: lower limit; UL: upper limit

Table A. 5 Risk of bias assessment by adapted Newcastle–Ottawa Scale for cross-sectional studies.

|  | Domains | **Selection (5 stars)** | | | | | **Comparability (2 stars)** | | **Outcome (3 stars)** | | | **Total score** |
| --- | --- | --- | --- | --- | --- | --- | --- | --- | --- | --- | --- | --- |
|  |  | Representativeness of the sample | Sample size | Non-respondents | Ascertainment of the exposure (risk factor) | The subjects in different outcome groups are comparable. based on the study design or analysis. Confounding factors are controlled | | Assessment of outcome | | Statistical test |  | |
| 1 | Ali. et al.. 2023 | 1 | 1 | 0 | 2 | 1 | | 2 | | 1 | 8 | |
| 2 | Cosemans. et al.. 2022 | 1 | 1 | 0 | 2 | 2 | | 2 | | 1 | 9 | |
| 3 | De Oliveira. et al.. 2019 | 0 | 1 | 0 | 1 | 0 | | 2 | | 1 | 5 | |
| 4 | De Prado-Bert. et al.. 2021 | 0 | 1 | 0 | 2 | 2 | | 2 | | 1 | 8 | |
| 5 | Dos Santos. et al.. 2022 | 0 | 1 | 0 | 1 | 1 | | 2 | | 1 | 6 | |
| 6 | Duan. et al.. 2017 | 0 | 1 | 0 | 0 | 1 | | 2 | | 1 | 5 | |
| 7 | Hoang. et al.. 2021 | 1 | 1 | 0 | 2 | 1 | | 2 | | 1 | 8 | |
| 8 | Kahl. et al.. 2018a | 0 | 1 | 0 | 1 | 1 | | 2 | | 1 | 6 | |
| 9 | Kahl. et al.. 2018b | 0 | 1 | 0 | 1 | 1 | | 2 | | 1 | 6 | |
| 10 | Kahl. et al.. 2018c | 0 | 1 | 0 | 1 | 1 | | 2 | | 1 | 6 | |
| 11 | Kahl. et al.. 2016 | 0 | 1 | 0 | 1 | 2 | | 2 | | 1 | 7 | |
| 12 | Karimi. et al.. 2020 | 1 | 1 | 0 | 2 | 1 | | 2 | | 1 | 8 | |
| 13 | Lucia. et al.. 2022 | 0 | 1 | 0 | 1 | 2 | | 2 | | 1 | 7 | |
| 14 | Lind. et al.. 2018 | 1 | 1 | 0 | 2 | 2 | | 2 | | 1 | 9 | |
| 15 | Ock. et al. (2020) | 1 | 1 | 0 | 2 | 2 | | 2 | | 1 | 9 | |
| 16 | Saad-Hussein. et al.. 2019 | 0 | 1 | 0 | 1 | 1 | | 2 | | 0 | 5 | |
| 17 | Shin. et al.. 2010 | 1 | 1 | 0 | 2 | 1 | | 2 | | 0 | 7 | |

Table A. 6 Risk of bias assessment by adapted Newcastle–Ottawa Quality Assessment Scale for cohort studies

|  | Domains | **Selection (4 stars)** | | | | **Comparability (2 stars)** | **Outcome (3 stars)** | | | **Total score** |
| --- | --- | --- | --- | --- | --- | --- | --- | --- | --- | --- |
|  |  | Representa-tiveness of the exposed cohort | Selection of the non-exposed cohort | Ascertainment of the exposure (risk factor) | Demonstration that outcome of interest was not present at start of study | Comparability of cohorts on the basis of the design or analysis | Assessment of outcome | Was follow-up long enough for outcomes to occur | Adequacy of follow up of cohorts |  |
| 1 | Andreotti et al.. 2015 | 1 | 0 | 1 | 1 | 1 | 1 | 1 | 1 | 7 |
| 2 | Guzzardi. et al.. 2016 | 1 | 0 | 1 | 1 | 2 | 1 | 1 | 1 | 8 |
| 3 | Hou. et al. (2013) | 1 | 0 | 1 | 1 | 2 | 1 | 1 | 1 | 8 |

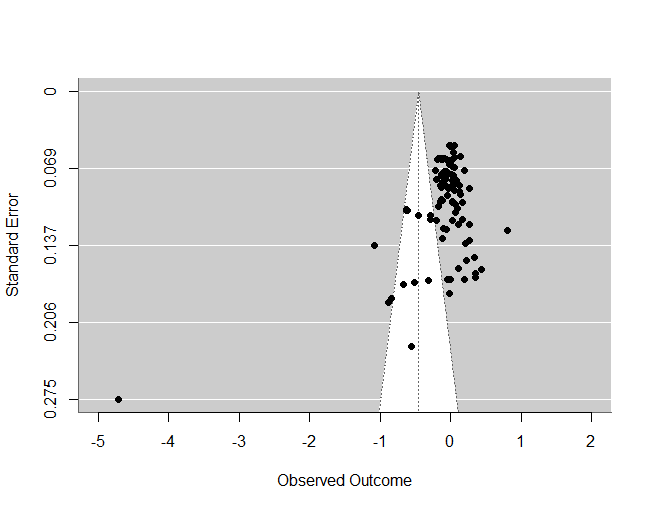

Fig. A.1 Funnel plot of associations between pesticide exposure and telomere length

Appendix B

(1) The formulas for calculating Hedges' g and transforming β coefficients from binary exposure variables into Hedges' g are as follows:

$$\begin{aligned} SMD=\frac{MD}{S_{pooled}}\#Eq.\left( B.1 \right) \end{aligned}$$

*MD* denotes the difference in mean telomere length between two independent groups, or the β coefficient from a linear regression model with a binary independent variable.

$$\begin{aligned} S_{pooled}=\sqrt{\frac{\left( n_{1}-1 \right)s_{1}^{2}+ \left( n_{2}-1 \right)s_{2}^{2}}{\left( n_{1}-1 \right)+ \left( n_{2}-1 \right)}}\#Eq.\left( B.2 \right) \end{aligned}$$

$S_{pooled}$ is the pooled standard deviation across two groups; $n_{1}$and $n_{2}$ are the sample size of two groups, and $s_{1}$ and $s_{2}$ are SD of two groups.

$$\begin{aligned} {SE}_{SMD}=\sqrt{\frac{n_{1}+n_{2}}{n_{1}n_{2}}+\frac{{SMD}^{2}}{2\left( n_{1}+n_{2} \right)}}\#Eq.\left( B.3 \right) \end{aligned}$$

$$\begin{aligned} g=SMD\times\left( 1-\frac{3}{4n-9} \right)\#Eq.\left( B.4 \right) \end{aligned}$$

$$\begin{aligned} {SE}_{g}=\sqrt{\frac{n_{1}+n_{2}}{n_{1}n_{2}}+\frac{g^{2}}{2\left( n_{1}+n_{2} \right)}}\#Eq.\left( B.5 \right) \end{aligned}$$

${SE}_{SMD}$, $g$ and ${SE}_{g}$ represent standard error of SMD, Hedges' g and standard error of Hedges' g.

(2) For studies reporting β coefficients from continuous exposure variables based on log-transformed values or percent change, the β coefficients are back-transformed to their untransformed values. The following equations 5-7 are then applied to transform β coefficients to partial correlations ($r_{p}$), while equations 8 and 9 are used to convert partial correlations to SMD (Ellis et al., 2023).

$$\begin{aligned} r_{p}=\frac{t}{\sqrt{t^{2}+df}}\#Eq.\left( B.6 \right) \end{aligned}$$

$$\begin{aligned} t=\frac{\beta}{{SE}_{\beta}}\#Eq.\left( B.7 \right) \end{aligned}$$

*t* represents the t-statistic of the β coefficient. If the study only reports p-value without SE or confidence interval, *t* is computed from *t* distribution table. *df* is the degree of freedom, calculated as df=n-p-1, where n and p are the total sample size and the number of predictors included in the model.

$$\begin{aligned} V_{rp}=\frac{\left( 1-r_{p}^{2} \right)^{2}}{df}\#Eq.\left( B.8 \right) \end{aligned}$$

$$\begin{aligned} SMD=\frac{2r_{p}}{\sqrt{1-r_{p}^{2}}}\#Eq.\left( B.9 \right) \end{aligned}$$

$$\begin{aligned} V_{SMD}=\frac{4V_{rp}}{\left( 1-r_{p}^{2} \right)^{3}}\#Eq.\left( B.10 \right) \end{aligned}$$

Where $V_{rp}$ and $V_{SMD}$ denotes the variance of $r_{p}$ and SMD.
